# Accurate prognosis for localized prostate cancer through coherent voting networks with multi-omic and clinical data

**DOI:** 10.1101/2022.07.28.22278156

**Authors:** Marco Pellegrini

## Abstract

**Background:** Prostate cancer is a very heterogeneous disease, from both a clinical and a biological/biochemical point of view, which makes the task of producing a stratification of patients into risk classes remarkably challenging. In particular, it is important an early detection and discrimination of the more indolent forms of the disease, from the more aggressive ones, requiring closer surveillance and timely treatment decisions.

**Methods:** We extend a recently developed supervised machine learning (ML) technique, called coherent voting networks (CVN) by incorporating novel model-selection technique to counter model overfitting. The CVN method is then applied to the problem of predicting an accurate prognosis (with a time granularity of 1 year) for patients affected by prostate cancer. The CVN is developed on a discovery cohort of 495 patients from the TCGA-PRAD collection, and validated on several other independent cohorts, comprising a gross total of 744 patients.

**Findings:** We uncover seven multi-gene fingerprints, each comprising six to seven genes, and a mixed clinical and genomic 5-marker fingerprint, that correspond to different input data types (clinical, mRNA expression, proteomic assays, methylation) and different time points, for the event of post-surgery progression-free survival (PFS) in patients diagnosed with prostate adenocarcinoma, who had not received prior treatment for their disease.

With a mixed 5-marker genomic and clinical fingerprint comprising Gleason primary score, tumor stage, psa, and molecular protein expression levels for CDKN1B and NF2 we attain on three independent cohorts statistically significant AUC values of 0.85, 0.88, and 0.87 respectively for PFS prediction at 3 years.

For purely genomic fingerprints, in seven independent cohorts for 21 combinations of cohort vs fingerprint, we report Odds Ratios ranging from a minimum of 9.0 and a maximum of 40.0, with average 17.5, geometric mean p-value 0.003; Cohen’s kappa values ranging from a minimum of 0.18 to a maximum of 0.65, with average 0.4; and AUC ranging from a minimum of 0.61 to a maximum of 0.88, with average 0.76, geometric mean p-value 0.001, for PFS prediction at 2, 3, and 4 years.

Many of the genes in our fingerprint have recorded prognostic power in some form of cancer, and have been studied for their functional roles in cancer on animal models or cell lines.

**Interpretation:** The development of novel ML techniques tailored to the problem of uncovering effective multi-gene prognostic biomarkers is a promising new line of attack for sharpening our capability to diversify and personalize cancer patient treatments. For the challenging problem of discriminating a fine time-scale for aggressive types of localized prostate cancer, we show that it is possible to attain more accurate prognostic predictions, with a granularity within a year, for the post-surgery early years.

## 1 Introduction

According to the World Cancer Research Fund International web site^1^, prostate cancer (PRC) is forecast as the second most commonly diagnosed cancer type in men (with 1.4 million cases worldwide) for the year 2022, and the 4th most commonly diagnosed cancer in the overall population (male and female).

The ECIS - European Cancer Information System^2^ predicts an incidence of 363,000 new diagnosed PRC for the EU27 + EFTA area in the year 2025 and estimates a mortality of about 78,000 due to PRC (representing about 10% of the deaths due to cancer in the male population, ranking third as cause of death by cancer type in the EU27+EFTA male population).

Siegel et al.^1^ report an estimate of 268,490 new cases of diagnosed prostate cancer for the year 2022 in the USA, and an estimate of 34,500 deaths due to prostate cancer (ranking second as cause of death by cancer type in the USA male population).

About 15% of the localized prostate cancer diagnoses are clinically classified as high risk^2^ and thus require timely management decisions. We concentrate our work on this fraction of the PRC patient population.

Prostate cancer is a very heterogeneous disease, from both a clinical and a biological/biochemical point of view, which makes the task of producing a stratification of patients into risk classes particularly challenging. In particular, it is important an early detection and discrimination of the indolent forms of the disease, from the aggressive ones, requiring closer surveillance^34^.

This report describes the application of a recently developed machine learning (ML) technique, called *coherent voting networks* to the problem of predicting an accurate prognosis for patients affected by prostate cancer (PRC).

Coherent voting networks (CVN) have been developed for the task of predicting overall survival (OS) of breast cancer (BC) patients at 5 years after surgery, based on tissue transcriptomic fingerprints (mRNA), and depending on the specific adjuvant therapy adopted^5^. Since CVN is a general ML technique it is natural to extend such technique to handling of further cancer types (in this report, prostate cancer), further data type (including clinical, miRNA, CNA, microbiome, methylation, proteomics, etc..), different time points and different events of interest. Moreover, in this report, we further develop the CVN technique in order to provide additional theoretical grounding to some of the algorithmic phases involved. Specifically, we re-examine the hyper-parameter optimization and feature selection phases (collectively indicated as model selection) and we show that a variation of the method by Andrew Ng^6^ to cope with model overfitting is both well grounded from a theoretical point of view and effective on our data.

We develop multi-gene fingerprints for predicting the risk of Progress-free survival (PFS) of patients over several time points. The molecular data sets for the fingerprint discovery are provided by the TCGA consortium and consist of assays of prostate biopsies and tissue removed via radical prostatectomy in patients diagnosed with prostate adenocarcinoma, who had not received prior treatment for their disease^3^.

Current diagnostic tests based on prostate-specific antigenes (PSA), Gleason score, Tumor stage, and other clinical measures often fail to distinguish between indolent and aggressive tumors, thus leading to over-diagnosis and over-treatment^78^910. This adverse phenomenon has been the driving force behind much recent research aiming at integrating PSA with molecular profiling or finding new alternative prognostic features leading to a more accurate PRC prognosis.

As of 2021, Manjang et al.^11^ list at least 32 prognostic genic signatures for PRC, however only a handful have been thoroughly validated, and made into commercially available kits (including Oncotype Dx^12^, Prolaris^13^, Decipher^14^, Decipher PORTOS^15^, and ProMark^16^). Such commercial kits are increasingly included in clinical protocols and practice^17^.

We focus on the task of proving a refined stratification and risk assessment for clinically classified high risk patients in the first five post-surgery years. Here we contribute to the search for effective multi-gene prognostic fingerprinting by applying the CVN to several omic and clinical data sets from prostate cancer patients to learn a pool of effective fingerprints. Next such fingerprints are applied to independent cohorts data sets to assess how performant these fingerprints can be (via a leave-one-out parameter optimization and bootstrapping performance evaluation). The reported results show remarkable promising performances in terms of Odds Ratio, Cohen’s kappa, and AUC, with good statistical significance. In particular fingerprint combining both clinical and ‘omic’ markers show encouraging results.

This paper is organized as follows. In Section 2 we report the main computational result on the performance of the proposed fingerprints. In Section 3 we recall the main steps of the CVN construction and usage, and we describe more in detail the novel model-selection techniques we introduce in this report. Finally, in Section 4 we comment on weak and strong points of our methodology and we place our work in the wider context of clinically useful prognostic tests for PRC.

## 2 Results

### 2.1 Clinical features of the discovery population

We use the TCGA-PRAD dataset for training validating and testing the prognostic CVN in the discovery phase and determining the best performing multi-gene fingerprints. We report in Table 1 the distributions of categorical attributes: progression-free survival status, tumor t-stage, tumor lymph node stage, radiation therapy, and reviewed Gleason sum.

**Table 1.** Categorical attributes of the TCGA PRAD patients. Progression free survival (PFS) event. 1=Progression, 0=Censored. Tumor stage. T34 includes T3A, T3B, T3C and T4. T2 includes T2A, T2B and T2C. Lymph Node Stage (American Joint Committee on Cancer Code). Reviewed Gleason Sum, LR (Low Risk) corresponds to levels 6 and 7, HR (High Risk) corresponds to levels 8,9 and 10.

|  | Train |  | Validation |  | Testing |  |
| --- | --- | --- | --- | --- | --- | --- |
| PFS event |  |  |  |  |  |  |
| Num | 122 | - | 64 | - | 55 | - |
| 0:CENSORED | 69 | 56% | 45 | 70% | 34 | 61% |
| 1:PROGRESSION | 53 | 43% | 19 | 29% | 21 | 38% |
| no data | 0 | - | 0 | - | 0 | - |
| Tumor stage |  |  |  |  |  |  |
| Num | 122 | - | 63 | - | 54 | - |
| T34 | 86 | 70% | 40 | 63% | 38 | 70% |
| T2 | 36 | 29% | 23 | 36% | 16 | 29% |
| no data | 0 | - | 1 | - | 1 | - |
| Lymph node stage |  |  |  |  |  |  |
| Num | 106 | - | 57 | - | 47 | - |
| N1 | 21 | 19% | 12 | 21% | 7 | 14% |
| N0 | 85 | 80% | 45 | 78% | 40 | 85% |
| no data | 16 | - | 7 | - | 8 | - |
| Radiation Therapy |  |  |  |  |  |  |
| Num | 119 | - | 62 | - | 54 | - |
| No | 100 | 84% | 56 | 90% | 48 | 88% |
| Yes | 19 | 15% | 6 | 9% | 6 | 11% |
| no data | 3 | - | 2 | - | 1 | - |
| Gleason sum |  |  |  |  |  |  |
| Num | 81 | - | 49 | - | 36 | - |
| LR | 55 | 67% | 34 | 69% | 27 | 75% |
| HR | 26 | 32% | 15 | 30% | 9 | 25% |
| no data | 41 | - | 15 | - | 19 | - |

We report in Table 2 the distributions of numerical attributes: progression-free survival timing, age at diagnosis, tumor mutation burden index, duration of follow-up, and PSA level before surgery,

**Table 2.**
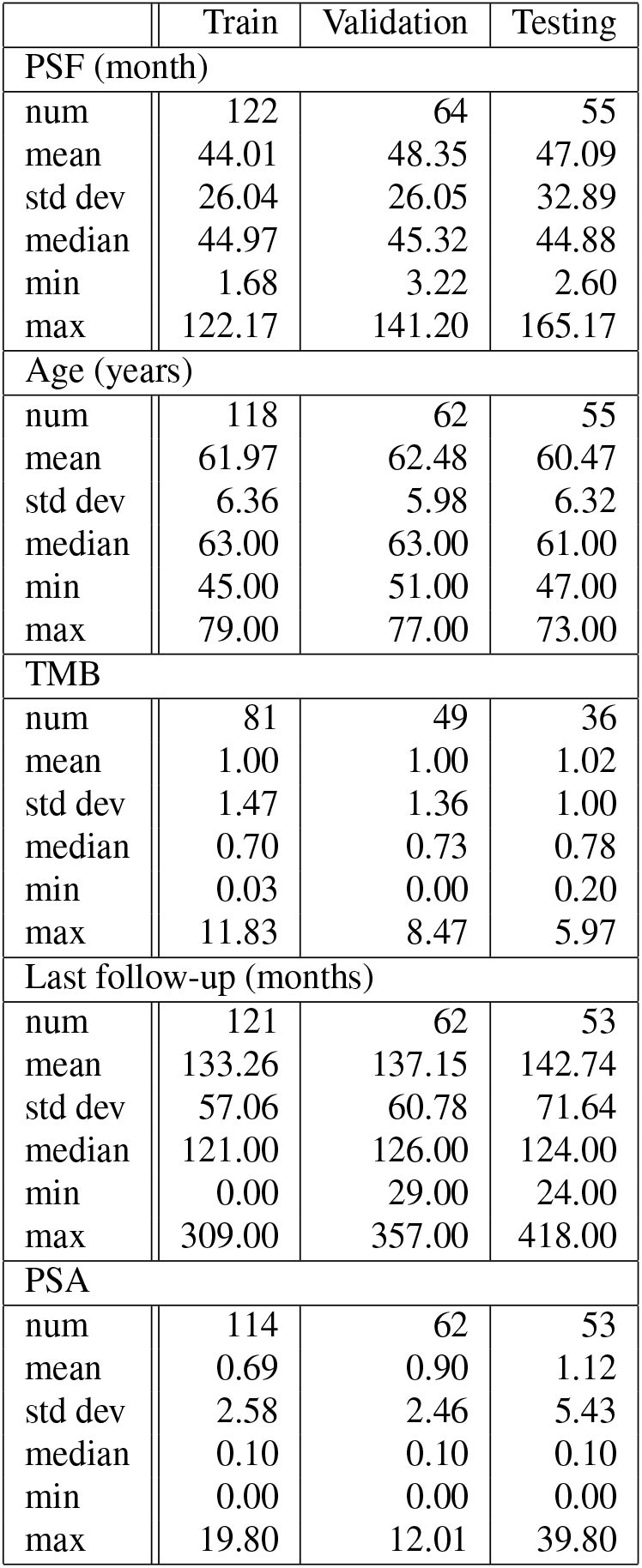
Numerical attributes of the TCGA PRAD patients. Progression free survival (PFS) time in months. Age at first diagnosis (years). Tumor mutation burden (TMB) nonsynonymous. Time interval from the date of initial pathologic diagnosis to the date of last followup (in months). Pre-operatory value of PSA.

|  | Train | Validation | Testing |
| --- | --- | --- | --- |
| PSF (month) |  |  |  |
| num | 122 | 64 | 55 |
| mean | 44.01 | 48.35 | 47.09 |
| std dev | 26.04 | 26.05 | 32.89 |
| median | 44.97 | 45.32 | 44.88 |
| min | 1.68 | 3.22 | 2.60 |
| max | 122.17 | 141.20 | 165.17 |
| Age (years) |  |  |  |
| num | 118 | 62 | 55 |
| mean | 61.97 | 62.48 | 60.47 |
| std dev | 6.36 | 5.98 | 6.32 |
| median | 63.00 | 63.00 | 61.00 |
| min | 45.00 | 51.00 | 47.00 |
| max | 79.00 | 77.00 | 73.00 |
| TMB |  |  |  |
| num | 81 | 49 | 36 |
| mean | 1.00 | 1.00 | 1.02 |
| std dev | 1.47 | 1.36 | 1.00 |
| median | 0.70 | 0.73 | 0.78 |
| min | 0.03 | 0.00 | 0.20 |
| max | 11.83 | 8.47 | 5.97 |
| Last follow-up (months) |  |  |  |
| num | 121 | 62 | 53 |
| mean | 133.26 | 137.15 | 142.74 |
| std dev | 57.06 | 60.78 | 71.64 |
| median | 121.00 | 126.00 | 124.00 |
| min | 0.00 | 29.00 | 24.00 |
| max | 309.00 | 357.00 | 418.00 |
| PSA |  |  |  |
| num | 114 | 62 | 53 |
| mean | 0.69 | 0.90 | 1.12 |
| std dev | 2.58 | 2.46 | 5.43 |
| median | 0.10 | 0.10 | 0.10 |
| min | 0.00 | 0.00 | 0.00 |
| max | 19.80 | 12.01 | 39.80 |

Overall, due to the randomized split of the patients, these features have similar distributions (mean, standard deviation) over the patient groups.

### 2.2 Performance on TCGA-PRAD data

In Table 3 we report seven fingerprints giving the best performance for different input data types (mRNA, proteomics, and methylation) and different time frames (year gap between high risk and low risk patients: 2-3, 3-4 and 4-5). For each fingerprint, the main measures of performance reported in Table 4 are odds ratio (OR), odds-ratio p-value, Cohen’s kappa, AUC, AUC p-value, and the log-rank test p-value. The odds ratios range from a minimum of 12.0 to a maximum of 21.0, with average 16.8, and all with significant p-values (except for fp14), geometric mean p-value 0.01. Cohen’s kappa ranges from a minimum of 0.29 to a maximum of 0.59, with average 0.47. AUC ranges from a minimum of 0.62 to a maximum 0.79, with average 0.72, with significant p-values (except for fp12) and geometric mean p-value 0.01. The log-rank p-values are all significant (except for fp14 which is borderline) and have geometric mean p-value 0.0006. Fingerprint fp14 has a significant AUC p-value, and fp12 has a significant OR p-value and log-rank p-value. Overall each fingerprint in Table 4 is statistically significant for at least one of the key measures. The Kaplan-Meier plots for these seven fingerprints are in Figures 1, 2, 3, 4, 5, 6, and 7, giving a graphical display of the good separation properties of the selected fingerprints. Additional performance measures including PPV/NPV and Sen/Spec are in the Github project repository.

**Table 3.** Listing of eight fingerprints with reference to time gap of high to low risk stratification in years, and to the omic data type. Genes are reported in HUGO nomenclature. Methylation loci are denoted with Illumina HumanMethylation450 BeadChip identification labels.

| Fp | Time | Data | genes | size |
| --- | --- | --- | --- | --- |
| Fp0 | 2-3 | mrna | CHST1 , GHRL , MAK , RAB11FIP4 , RPEL1 , ZEB1 | 6 |
| Fp1 | 3-4 | mrna | ASH1L-AS1 , C1orf88 , DBN1 , HRSP12 , MAFG , SNORA18 , TRIM65 | 7 |
| Fp12 | 2-3 | rppa | CDKN1B , MAPK9 , MYC , NDRG1 , NF2 , RB1 , SCD | 7 |
| Fp14 | 4-5 | rppa | CDH1 , DIABLO , EGFR , GAB2 , PRKCA , RPS6KB1 | 6 |
| Fp30 | 3-4 | rppa | CDK1 , CDKN1B , CLDN7 , MYC , NF2 , SCD | 6 |
| Fp20 | 2-3 | mrna+rppa | BAK1 , PTCHD4 , FANCC , FBRSL1 , OMP , SULT1C3 , CDKN1B | 7 |
| Fp37 | 3-4 | methyl | cg02928644, cg03062002, cg11504897, cg11620238, cg22337128, cg22661239 | 6 |
| Fp160 | 3-4 | clinical+rppa | PSA, Tumor Stage, Gleason primary score, NF2, CDKN1B | 5 |

**Table 4.**
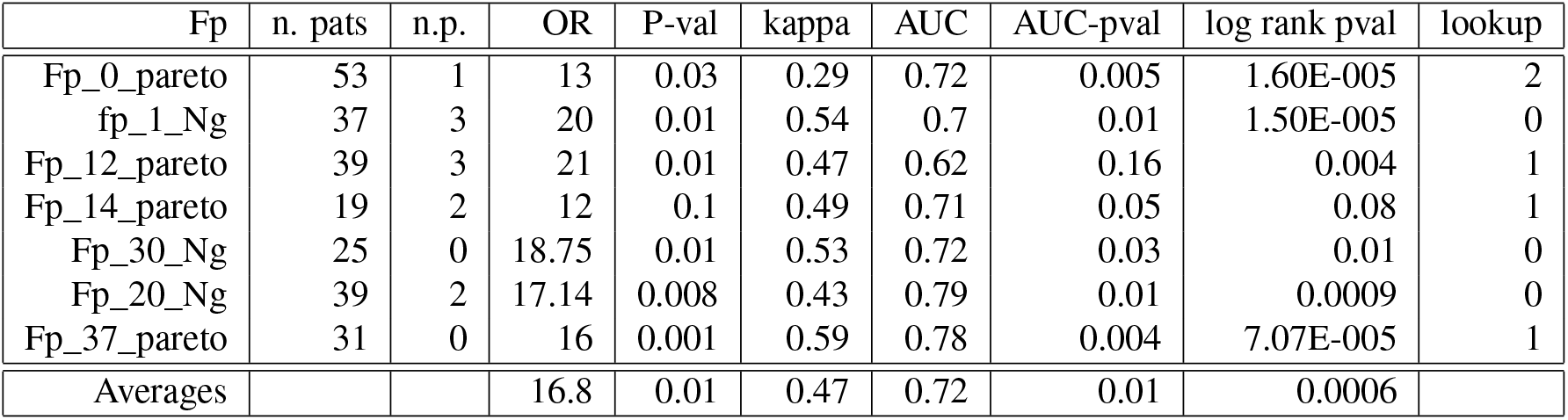
Performance measures of seven fingerprints (Fp) on TCGA-PRAD discovery data. The performance is measured on the test data after training (on train data) and model selection (on validation data). We report the whether the fingerprint has bee selected via Pareto-based or Ng-based model selection. The table reports the fingerprint identifier (Fp), the number of patients in the test set (n. pats), the number of no predictions (n.p.), the odds ratio (OR), its p-value, the Cohen’s kappa value, the area under the curve (AUC), its p-value (AUC-pval), the p-value of the log-rank test, and the lookup number. For Ng-based model selection that does not use lookup, the lookup number is set to 0 by default. For averaging p-values we use the geometric mean, for other values the arithmetic mean. Results in this table are obtained with sw pipeline (a).

| Fp | n. pats | n.p. | OR | P-val | kappa | AUC | AUC-pval | log rank pval | lookup |
| --- | --- | --- | --- | --- | --- | --- | --- | --- | --- |
| Fp_0_pareto | 53 | 1 | 13 | 0.03 | 0.29 | 0.72 | 0.005 | 1.60E-005 | 2 |
| fp_1_Ng | 37 | 3 | 20 | 0.01 | 0.54 | 0.7 | 0.01 | 1.50E-005 | 0 |
| Fp_12_pareto | 39 | 3 | 21 | 0.01 | 0.47 | 0.62 | 0.16 | 0.004 | 1 |
| Fp_14_pareto | 19 | 2 | 12 | 0.1 | 0.49 | 0.71 | 0.05 | 0.08 | 1 |
| Fp_30_Ng | 25 | 0 | 18.75 | 0.01 | 0.53 | 0.72 | 0.03 | 0.01 | 0 |
| Fp_20_Ng | 39 | 2 | 17.14 | 0.008 | 0.43 | 0.79 | 0.01 | 0.0009 | 0 |
| Fp_37_pareto | 31 | 0 | 16 | 0.001 | 0.59 | 0.78 | 0.004 | 7.07E-005 | 1 |
| Averages |  |  | 16.8 | 0.01 | 0.47 | 0.72 | 0.01 | 0.0006 |  |

**Figure 1.**
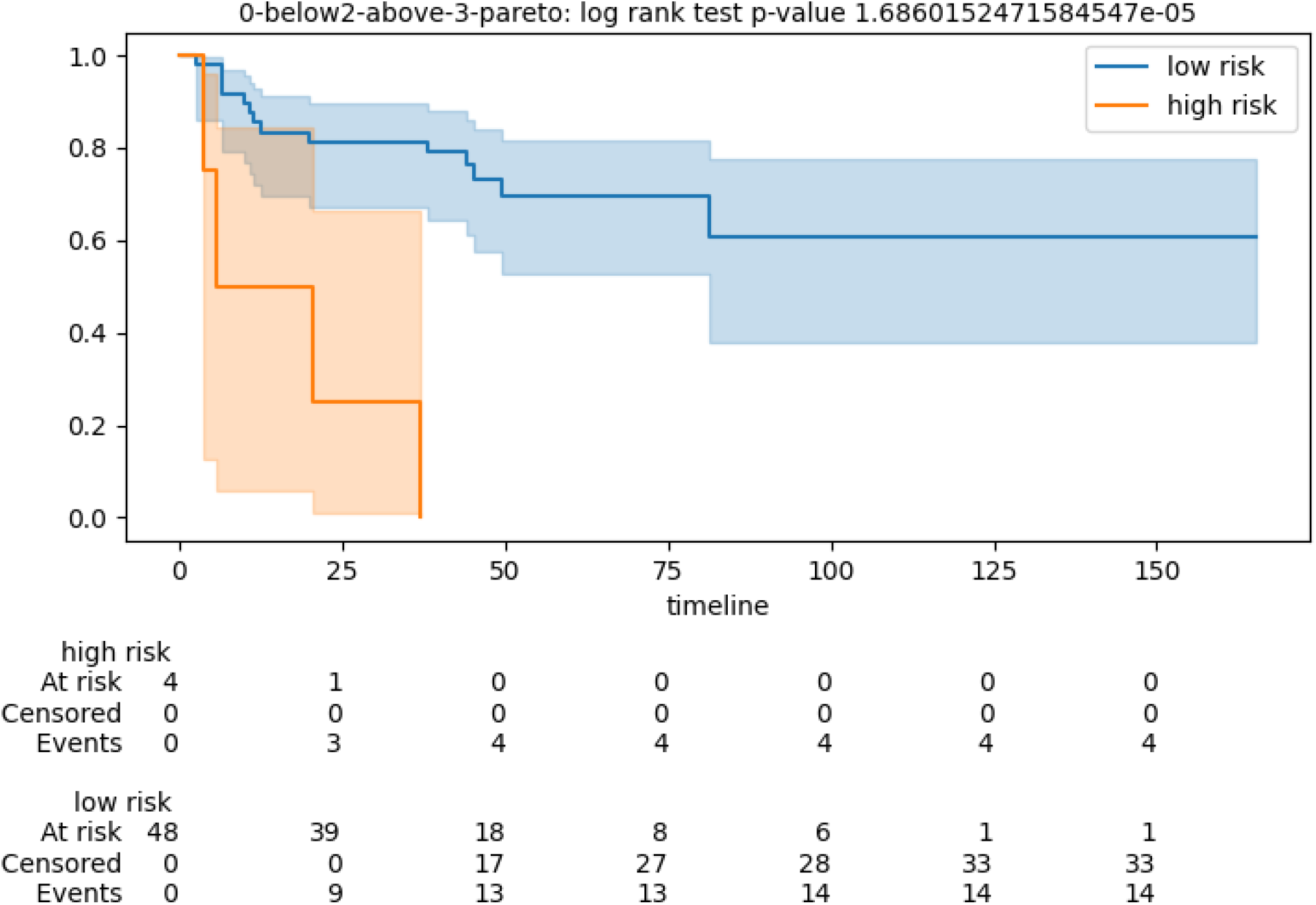
Kaplan-Meier for test cohort stratification of patients according to the CVN with fingerprint fp0. Low risk category corresponds to patients experiencing disease progression after 3 years. High risk category corresponds to patients experiencing disease progression before 2 years. Gene expression is measured via mRNA expression levels. Timeline in months.

**Figure 2.**
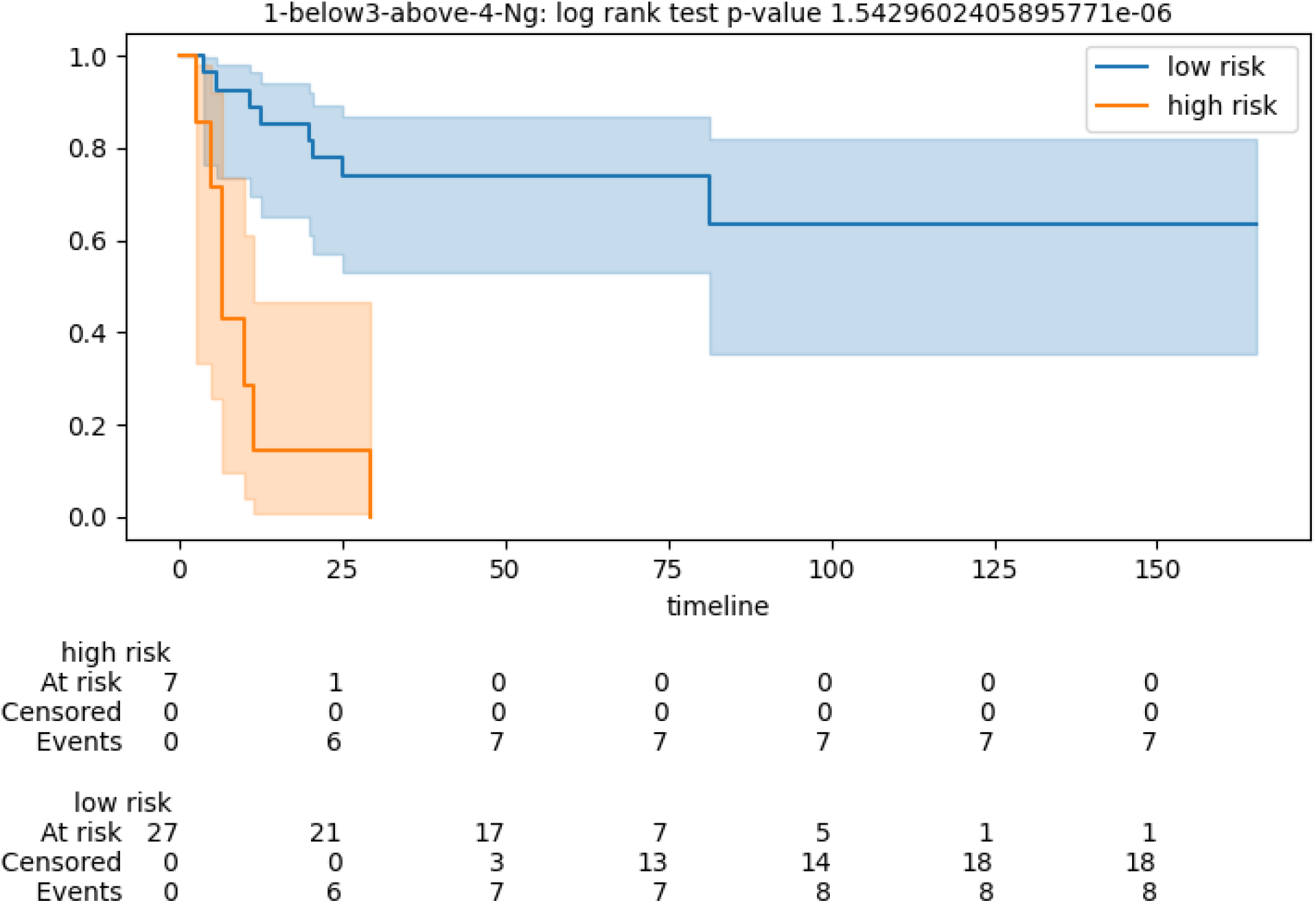
Kaplan-Meier for test cohort stratification of patients according to the CVN with fingerprint fp1. Low risk category corresponds to patients experiencing disease progression after 4 years. High risk category corresponds to patients experiencing disease progression before 3 years. Gene expression is measured via mRNA expression levels. Timeline in months.

**Figure 3.**
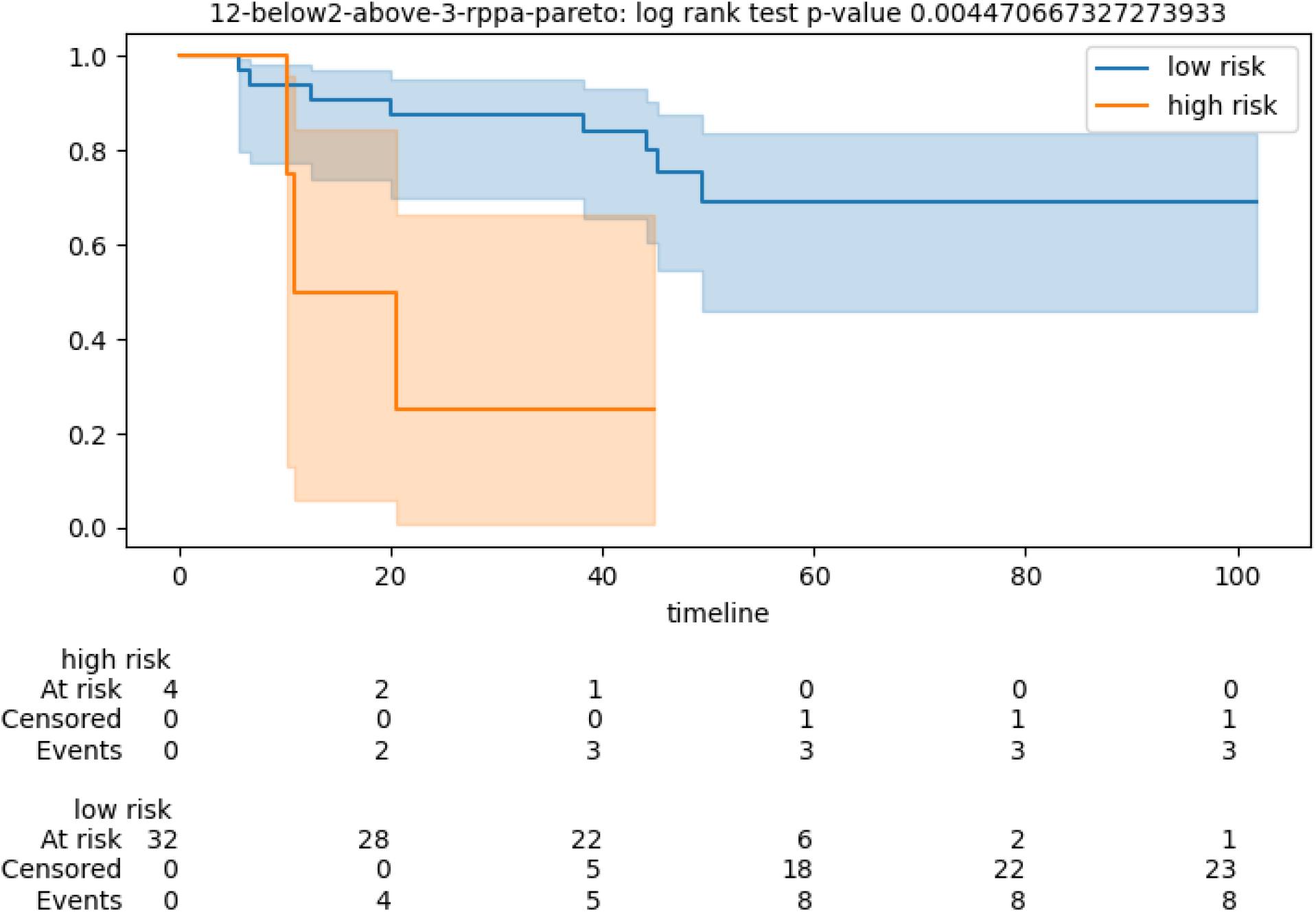
Kaplan-Meier for test cohort stratification of patients according to the CVN with fingerprint fp12. Low risk category corresponds to patients experiencing disease progression after 3 years. High risk category corresponds to patients experiencing disease progression before 2 years. Protein expression is measured via rppa assay. Timeline in months.

**Figure 4.**
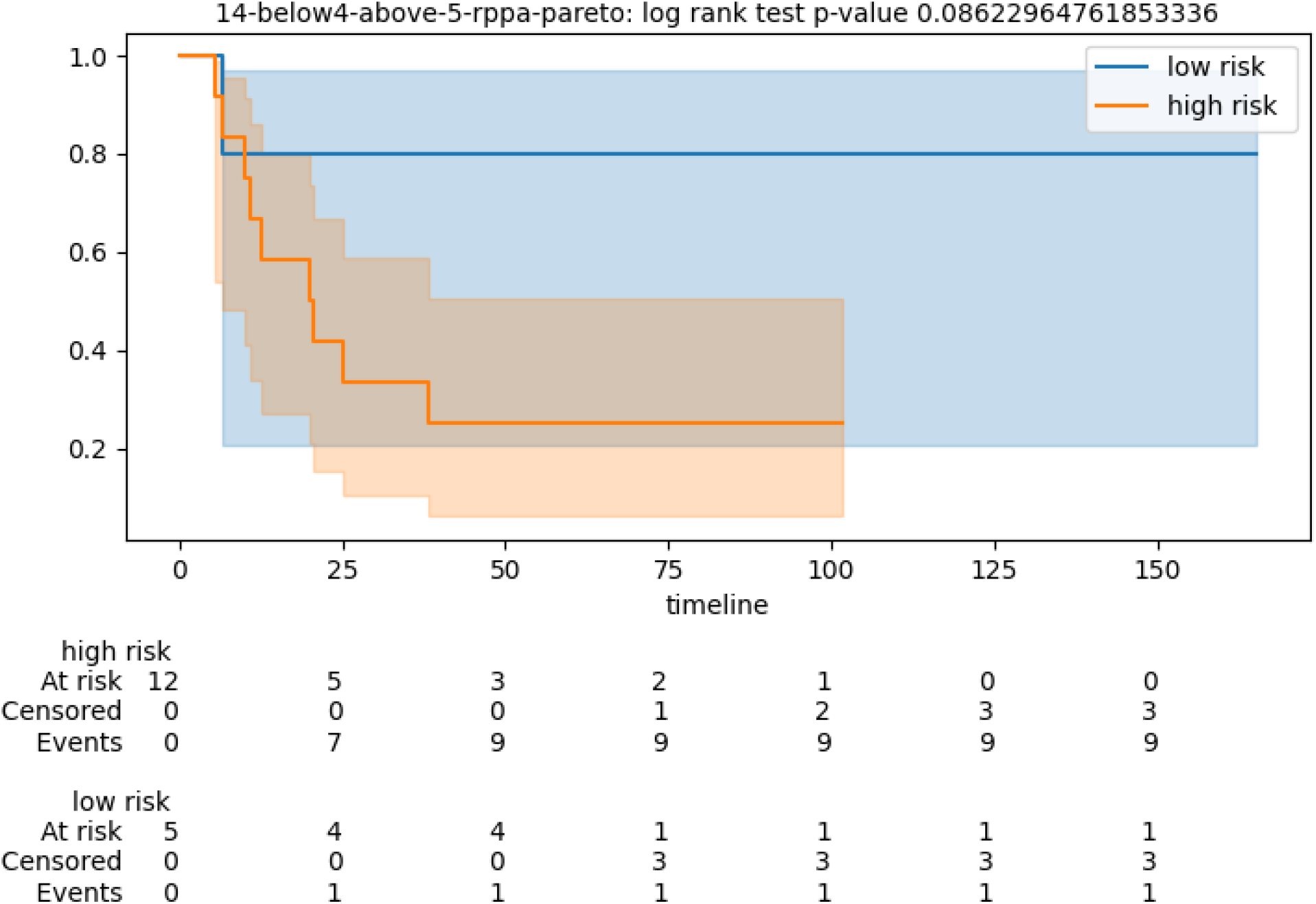
Kaplan-Meier for test cohort stratification of patients according to the CVN with fingerprint fp14. Low risk category corresponds to patients experiencing disease progression after 5 years. High risk category corresponds to patients experiencing disease progression before 4 years. Protein expression is measured via rppa assay. Timeline in months.

**Figure 5.**
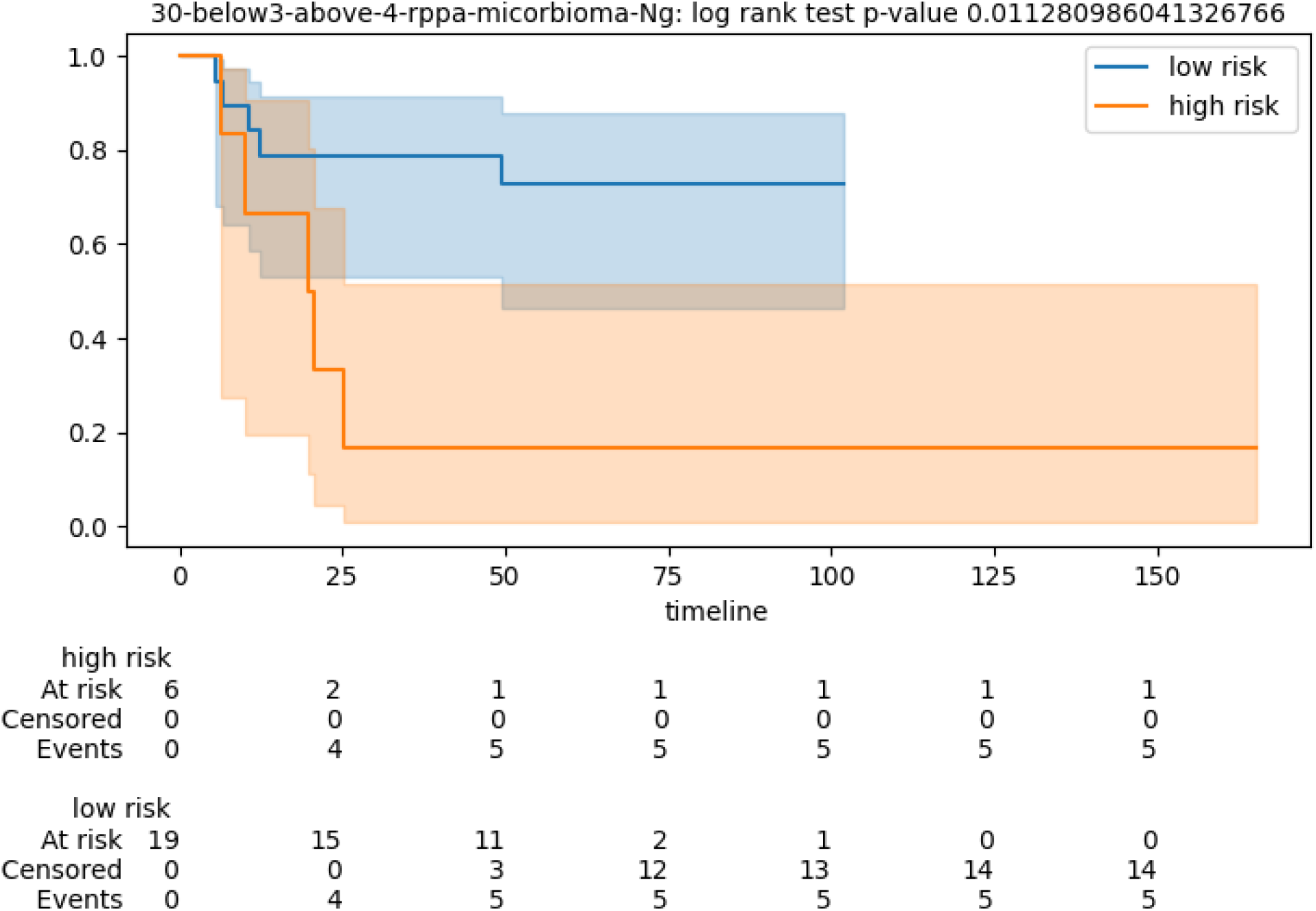
Kaplan-Meier for test cohort stratification of patients according to the CVN with fingerprint fp30. Low risk category corresponds to patients experiencing disease progression after 4 years. High risk category corresponds to patients experiencing disease progression before 3 years. Protein expression is measured via rppa assay. Timeline in months.

**Figure 6.**
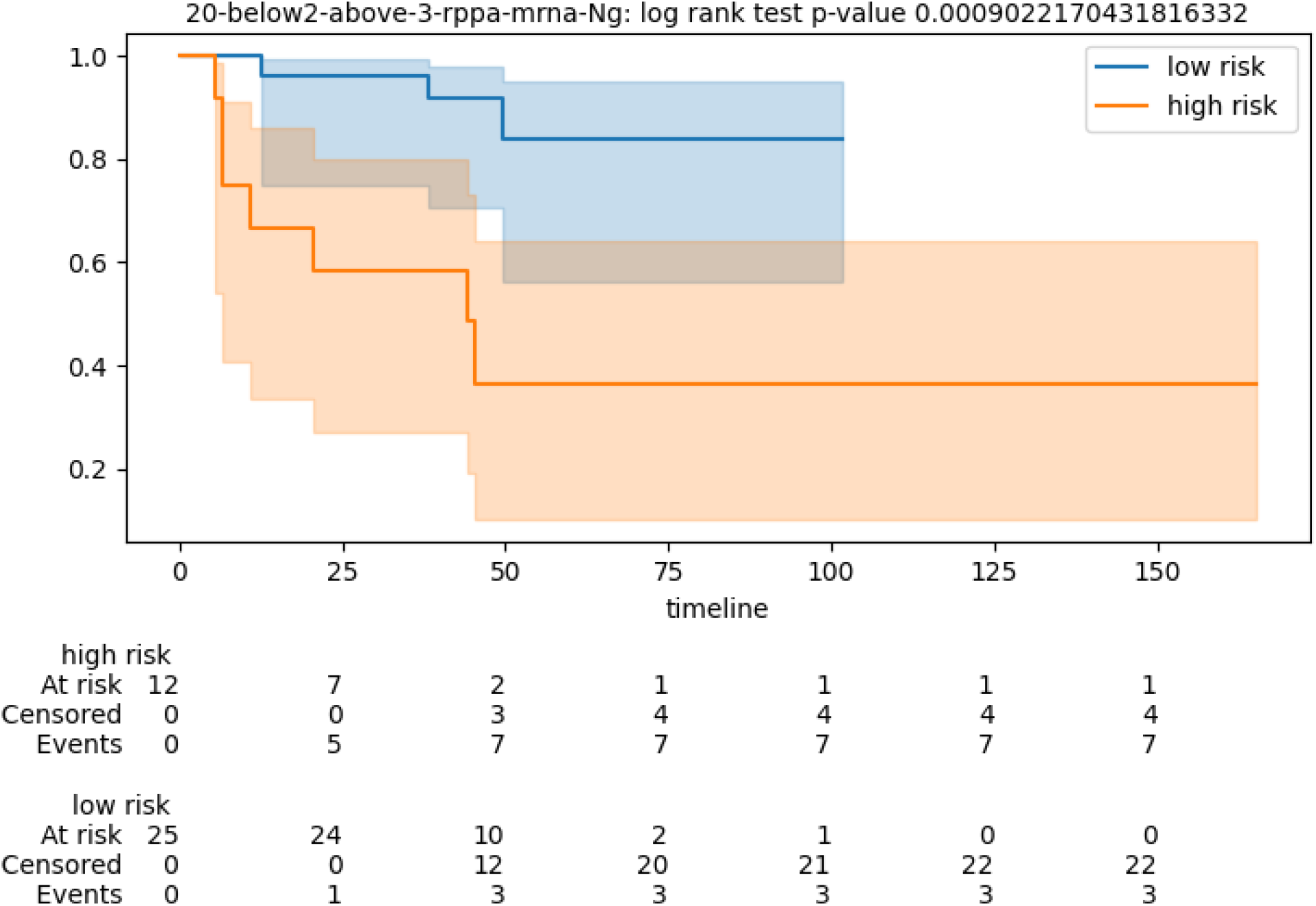
Kaplan-Meier for test cohort stratification of patients according to the CVN with fingerprint fp20. Low risk category corresponds to patients experiencing disease progression after 3 years. High risk category corresponds to patients experiencing disease progression before 2 years. The fingerprint is mixed. Protein expression is measured via rppa assay. Gene expression levels are measured via mRNA expression levels. Timeline in months.

**Figure 7.**
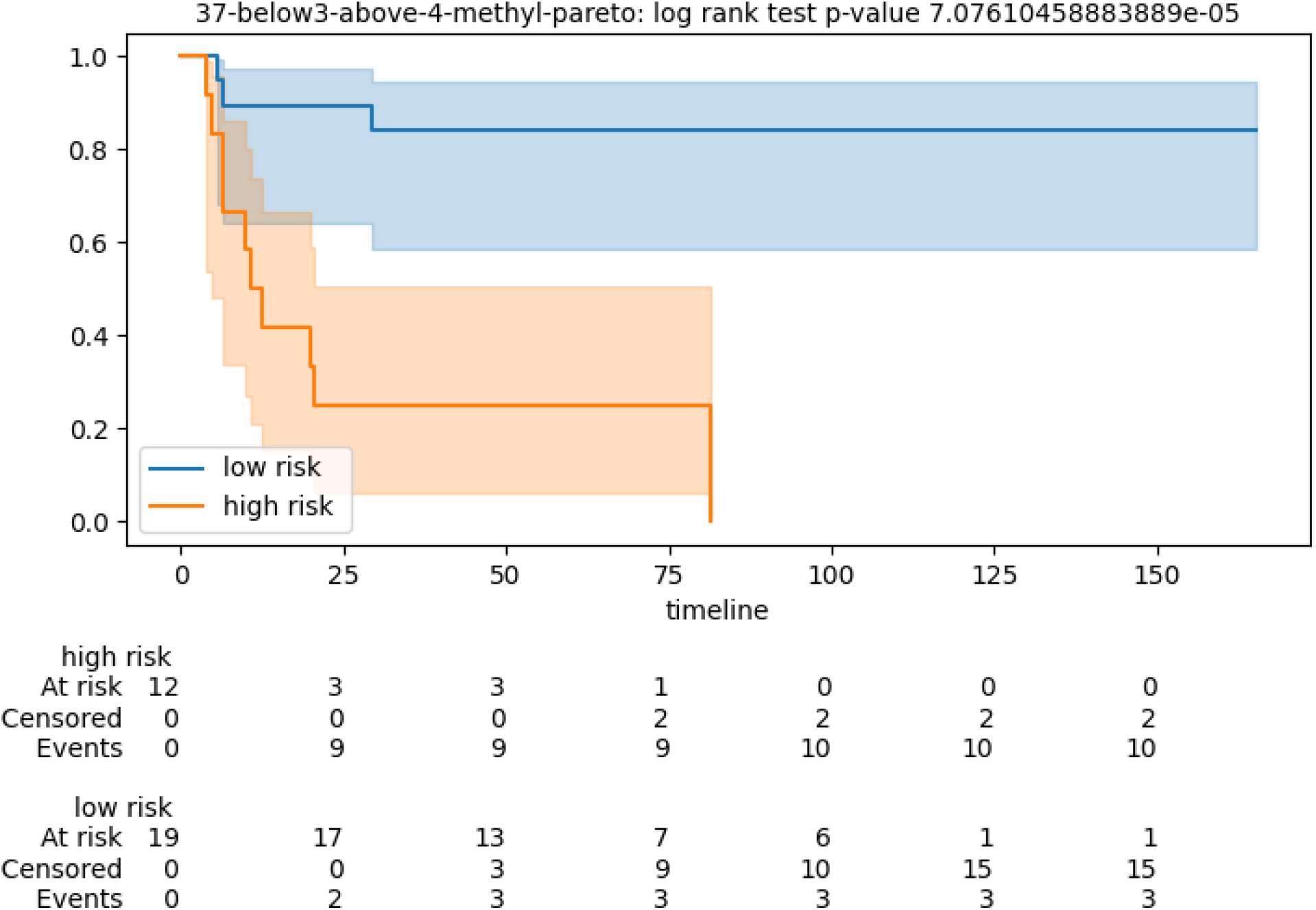
Kaplan-Meier for test cohort stratification of patients according to the CVN with fingerprint fp37. Low risk category corresponds to patients experiencing disease progression after 4 years. High risk category corresponds to patients experiencing disease progression before 3 years. Fingerprint based on a measure of the methylation level of methylation loci. Timeline in months.

### 2.3 Independent cohorts

In order to validate the selected fingerprints, we measure their prognostic performance on seven independent cohorts of PRC patients (listed in Table 9) with a raw total of 744 patients. These independent data sets have been produced with several platforms and include as event endpoints: Overall survival (OS), Biochemical recurrence (BCR), Disease-free survival (DFS), or a category-based High-risk/Low-risk assessment. On these independent cohorts, we fix the gene fingerprint and we generate predictors for leave-one-out (LOO) assays on the full range of hyperparameters for CVN, finally selecting the best performing configuration in terms of OR (or Cohen’s kappa), subject to a limit on the number of no predictions below 15%. Since it is known that leave-one-out cross-validation has a low bias but a high variance in performance estimation of the generalization error, we perform a bootstrap performance estimation of the selected configuration (fingerprint plus hyperparameters) using a theory of Efron and Tibshirani (1997)^18^ (more details in the Methods section). Table 5 reports the combinations of data sets and fingerprints for which we obtain OR at least 8.0, and no prediction below 20% of the number of patients in the bootstrapping assay. All results reported are statistically significant (below 0.05) in p-value for at least one key measure (OR or AUC). The Odds ratio ranges from a minimum of 8.33 to a maximum of 40.0 with average 17.5 and (geometric) mean p-value 0.003. Cohen’s kappa ranges from a minimum of 0.18 to a maximum of 0.65 with average 0.4, while AUC values range from 0.61 to 0.88, with average 0.76 and (geometric) mean p-value 0.001. Interestingly, the best performance in terms of OR and kappa is attained for fp1 on data set GSE46602 which is the most balanced data set in our pool having about 50% of high-risk and 50% low-risk patients. The selected fingerprint appears to have good prognostic performance across a wide choice of molecular measurement platforms, event end-points, and patients’ clinical conditions.

**Table 5.** Performance of the seven CVN selected fingerprints over seven independent cohorts of PRC patients using loo model selection and bootstrap performance evaluation. We report 21 combinations of cohort vs fingerprints attaining OR above 8.0. The table lists the independent cohort id (Dataset), the fingerprint ID (Fp), the number of patients (n. pats), the average number of no predictions (n.p.), the estimation of the Odds ratio (OR), its p-value (P-val), and the estimation of Cohen’s kappa (kappa), based on the expected values of true/false positives and true/false negatives by the bootstrapping. The area under the curve (AUC) value and its p-value (AUC-pval) are measured for the consensus predictor obtained by the bootstrapping. GSE84042 is the only independent data set in our pool with methylation data fit for validating fp37. Proteomic fingerprints have been validated on mRNA data of independent cohorts. For averaging p-values we use the geometric mean, for other values the arithmetic mean. Results in this table are obtained with sw pipeline (b).

| Dataset | Fp | n. pats | n.p. | OR | P-val | kappa | AUC | AUC-pval |
| --- | --- | --- | --- | --- | --- | --- | --- | --- |
| MSKCC | Fp_0 | 110 | 1.7 | 15.4 | 0.0001 | 0.4 | 0.68 | 0.003 |
| MSKCC | Fp_0 | 93 | 3.77 | 13.11 | 0.0007 | 0.32 | 0.7 | 0.0006 |
| GSE70769 | Fp_0 | 39 | 10.1 | 12 | 0.02 | 0.42 | 0.77 | 0.007 |
| GSE46602 | Fp_0 | 30 | 1.2 | 9.2 | 0.06 | 0.33 | 0.72 | 0.01 |
| GSE53922 | Fp_0 | 89 | 14.32 | 8.33 | 0.01 | 0.26 | 0.64 | 0.02 |
| GSE46602 | Fp_14 | 30 | 5.06 | 9 | 0.03 | 0.43 | 0.78 | 0.005 |
| GSE53922 | Fp_14 | 90 | 26.08 | 20 | 0.003 | 0.43 | 0.82 | 0.0001 |
| MSKCC | Fp_1 | 110 | 3.9 | 8.66 | 0.0007 | 0.37 | 0.69 | 9.70E-005 |
| GSE46602 | Fp_1 | 30 | 4.05 | 30 | 0.002 | 0.59 | 0.86 | 0.0003 |
| GSE46602 | Fp_1 | 30 | 1.69 | 40 | 0.0005 | 0.65 | 0.83 | 0.0009 |
| GSE46602 | Fp_12 | 30 | 3.7 | 25 | 0.003 | 0.59 | 0.86 | 0.0006 |
| GSE84042 | Fp_12 | 65 | 7.3 | 18.33 | 0.12 | 0.37 | 0.88 | 0.002 |
| MSKCC | Fp_30 | 110 | 2.36 | 13.83 | 0.02 | 0.18 | 0.61 | 0.04 |
| GSE54460 | Fp_30 | 31 | 1.47 | 20 | 0.16 | 0.48 | 0.79 | 0.05 |
| GSE46602 | Fp_30 | 30 | 5.3 | 13.5 | 0.01 | 0.47 | 0.82 | 0.002 |
| GSE53922 | Fp_30 | 89 | 7.66 | 12 | 0.004 | 0.21 | 0.7 | 0.001 |
| MSKCC | Fp_20 | 93 | 2.2 | 22.47 | 0.0006 | 0.32 | 0.7 | 0.001 |
| GSE46602 | Fp_20 | 30 | 0.5 | 30 | 0.001 | 0.55 | 0.83 | 0.001 |
| GSE53922 | Fp_20 | 89 | 40 | 16 | 0.002 | 0.43 | 0.79 | 0.0007 |
| GSE37199 | Fp_20 | 107 | 28.2 | 10.88 | 0.001 | 0.33 | 0.69 | 0.001 |
| GSE84042 | Fp_37 | 145 | 20.4 | 19.9 | 0.000017 | 0.46 | 0.87 | 1.90E-006 |
| Averages |  |  |  | 17.5 | 0.003 | 0.4 | 0.76 | 0.001 |

Of the seven independent cohorts in our assessment, five are obtained via surgically removed tumor tissues (through either biopsies or radical prostatectomy), thus consistent with the specimens used in the discovery cohort. Two independent cohorts (GSE37199 and GSE53922) are based instead on blood samples of PRC patients. Unexpectedly, fingerprint Fp20 has significant discriminative power also on both of these test cases, however the number of no predictions increases to 30-40% of the patient cohort.

### 2.4 Mixed clinical and genomic fingerprints

From the TCGA PRAD clinical data file we have selected 24 clinical/pathological features known to have prognostic power in prostate cancer. We have appended these features to the omic molecular expression matrices and iterated the fingerprint discovery pipeline. The fingerprint fp160 composed of three clinical parameters: Gleason primary score, tumor stage, psa, and two molecular protein expression levels for CDKN1B and NF2 has emerged as very concise and performant. Performance measures are reported in Table 6 both for the discovery pipeline and for the validation pipeline on independent cohorts. Figure 8 is the corresponding Kaplan-Meier plot. The AUC measure is 0.87 at p-value 0.001 on TCGA test data and is consistently confirmed in three independent cohorts bootstrap evaluations as well as in bootstrapping on the complete TCGA cohort. This mixed fingerprint attains better performance (in terms of AUC) with respect to the predictors obtained using the same discovery procedure starting from the 24 clinical/pathological features alone, or from the genomic data alone (data not shown). The mixed fingerprint has also better performace than a fingerprint composed of Gleason total score, tumor stage, psa, and age (data not shown).

**Table 6.** Performance of the mixed clinical and molecular fingerprint fp160 on the TCGA PRAD rppa data set and on independent cohorts. In the notes, it is reported the software pipeline used, and whether the input data set has been equalized with a size ratio of the two labels up to 3-to-1. n.p. is the number of no predictions. The year gap is between 3 and 4.

| Dataset | fp type | n. patients | n.p. | or | or pval | kappa | auc | auc pval | notes |
| --- | --- | --- | --- | --- | --- | --- | --- | --- | --- |
| TCGA-rppa | fp160 | 25 | 3 | 18 | 0.008 | 0.6 | 0.87 | 0.001 | sw (a) |
| TCGA-rppa | fp160 | 126 | 22 | 18.8 | 2.1E-10 | 0.61 | 0.83 | 4.8E-9 | sw (b) |
| GSE46602 | fp160 | 30 | 0 | 28.8 | 0.0007 | 0.6 | 0.85 | 0.0006 | sw (b), eq |
| GSE70769 | fp160 | 20 | 2 | 39.0 | 0.01 | 0.67 | 0.88 | 0.01 | sw (b), eq |
| GSE84042 | fp160 | 36 | 6 | 22.0 | 0.007 | 0.58 | 0.87 | 0.002 | sw (b),eq |

**Figure 8.**
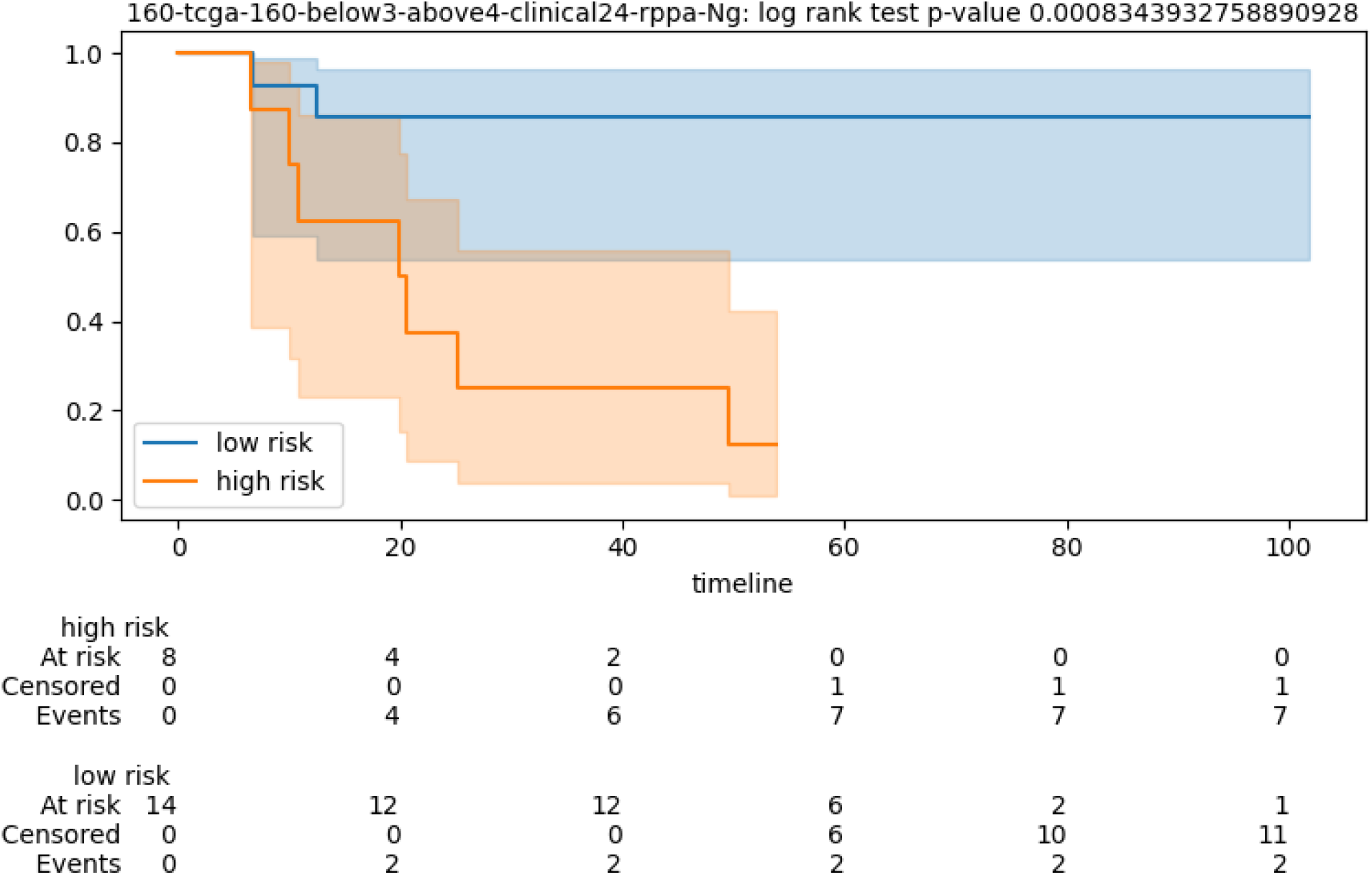
Kaplan-Meier for test cohort stratification of patients according to the CVN with fingerprint fp160. Low risk category corresponds to patients experiencing disease progression after 4 years. High risk category corresponds to patients experiencing disease progression before 3 years. Fingerprint based on mixed clinical and onic measurements. Timeline in months.

### 2.5 Biological relevance of the genes in the fingerprints

In this section, we report the outcome of investigating the role of each gene of our multi-gene fingerprints in the progress of prostate cancer. We could not find evidence that significant subsets of the genes in our fingerprints have been analyzed together previously in the context of prostate cancer. First, we use two established online databases to explore the prognostic power and the oncological annotations of each gene separately with respect to cancer in general. Next, we do a literature search of articles reporting on direct or indirect functional associations of each gene to prostate cancer (or to solid tumors, more widely).

#### 2.5.1 Human Protein Atlas and COSMIC

Searches of the protein-coding genes from our fingerprints in The Human Protein Atlas database^4^ show evidence of some prognostic power (relative to eventual OS) for 29 out of 37 for some cancer types (mostly kidney) (see Table 11). This database records only one gene of our pool as having prognostic power in prostate cancer (for eventual OS). We notice however that a recent study^19^ on the TCGA data quality for survival analysis indicates that the TCGA-PRAD OS annotations may be deficient, due to relatively short follow-up, thus the lack of prognostic power for each individual gene in prostate cancer may be explained. TCGA-PRAD records for PFS are of good quality, in contrast.

**Table 7.** Performance of fingerprints and corresponding bootstrap consensus predictors on subsets of patients identified as High risk (HR) or Intermediate Risk (IR) by two stratification schemes based on tumor stage, PSA and Gleason score: the D’Amico scheme^49^ and the NICE scheme^50^. The table reports the fingerprint identifier (Fp), the independent cohort identifier (Dataset), the time gap of high to low risk stratification in years for the CVN method (Time) the number of patients in the subset of patients (n. pats), the number of no predictions (n.p.), the odds ratio (OR), its p-value (P-val), and the Cohen’s kappa (kappa) for the bootstrap consensus predictor on the subset of patients.

| fp | Dataset | Time | stratum | n. pats | n.p. | OR | P-val | kappa |
| --- | --- | --- | --- | --- | --- | --- | --- | --- |
| fp37 | GSE84042 | 4-5 | nice IR | 48 | 4 | 9.3 | 0.04 | 0.29 |
| fp37 | GSE84042 | 4-5 | nice HR | 41 | 9 | 27 | 0.0003 | 0.66 |
| fp37 | GSE84042 | 4-5 | damico HR | 79 | 11 | 20.5 | 4.00E-005 | 0.5 |
| fp0 | GSE46602 | 2-3 | damico HR | 28 | 1 | 8.12 | 0.07 | 0.31 |
| fp0 | GSE46602 | 2-3 | nice HR | 20 | 1 | 4.2 | 0.03 | 0.23 |
| fp1 | GSE46602 | 2-3 | damico HR | 28 | 0 | 36 | 0.0004 | 0.71 |
| fp1 | GSE46602 | 2-3 | nice IR | 9 | 0 | 7 | 0.37 | 0.6 |
| fp1 | GSE46602 | 2-3 | nice HR | 20 | 0 | 33 | 0.004 | 0.68 |
| fp12 | GSE46602 | 2-3 | damico HR | 28 | 2 | 66 | 0.0001 | 0.76 |
| fp12 | GSE46602 | 2-3 | nice IR | 9 | 2 | 5 | 0.46 | 0.58 |
| fp12 | GSE46602 | 2-3 | nice HR | 20 | 0 | 38.5 | 0.002 | 0.79 |
| fp20 | GSE46602 | 3-4 | damico HR | 28 | 0 | 20 | 0.003 | 0.61 |
| fp20 | GSE46602 | 3-4 | nice HR | 22 | 0 | 14 | 0.02 | 0.54 |
| fp1 | GSE46602 | 3-4 | damico HR | 28 | 0 | 28 | 0.001 | 0.6 |
| fp1 | GSE46602 | 3-4 | nice HR | 22 | 0 | 30 | 0.009 | 0.63 |
| fp14 | GSE46602 | 3-4 | damico HR | 28 | 2 | 17.3 | 0.003 | 0.6 |
| fp14 | GSE46602 | 3-4 | nice HR | 22 | 2 | 12 | 0.03 | 0.52 |
| fp0 | GSE70769 | 2-3 | damico HR | 17 | 2 | 26 | 0.06 | 1 |
| fp0 | GSE70769 | 2-3 | nice IR | 22 | 3 | 11 | 0.1 | 0.53 |
| fp0 | GSE70769 | 2-3 | nice HR | 16 | 2 | 13 | 0.24 | 1 |

**Table 8.** Performance evaluation of randomly generated fingerprints. The random fingerprint size is fixed equal to the size of the corresponding fingerprint in Table 3. The random sampling is performed on the genes passing the initial statistical filter. The table reports the performance of the model with best OR among the Pareto-based and the Ng-based selected models. The table reports the input file ID (file n.) corresponding to the seven fingerprints in Table 3, the fixed size of the sampled fingerprints (fp size), the number of patients in the test set (n. pats), the number of no predictions (n.p.), the odds ratio (OR), its p-value (P-val), the Cohen’s kappa (kappa), the area under the curve (AUC) value, its p-value (AUC-pval), and the lookup number. The lookup number is default 0 for models Ng-based.

| file n. | fp size | n. pats | n.p. | OR | P-val | kappa | AUC | AUC-pval | lookup |
| --- | --- | --- | --- | --- | --- | --- | --- | --- | --- |
| 0 | 6 | 53 | 0 | 4 | 0.08 | 0.23 | 0.65 | 0.04 | 0 |
| 1 | 7 | 37 | 0 | 11.3 | 0.003 | 0.47 | 0.79 | 0.001 | 3 |
| 12 | 7 | 39 | 0 | 9.33 | 0.04 | 0.4 | 0.83 | 0.004 | 2 |
| 14 | 6 | 19 | 0 | 1.5 | 1 | 0.09 | 0.65 | 0.14 | 0 |
| 30 | 6 | 25 | 4 | 14 | 0.05 | 0.48 | 0.74 | 0.04 | 2 |
| 20 | 7 | 39 | 2 | 8.66 | 0.02 | 0.41 | 0.7 | 0.03 | 2 |
| 37 | 6 | 31 | 4 | 0.3 | 0.6 | -0.17 | 0.49 | 0.54 | 0 |

**Table 9.** Technological platforms for measuring molecular species in the discovery cohort (TCGA-PRAD) and in the independent cohorts. We report the platforms corresponding to the molecular data (mRNA, rppa, methylation) used in this study. The table lists the cohort identifier (ID), the end point event (E.P.), the technological platforms (Platform), and the raw number of patients of the cohort (n. pats). Number of patients refers to the raw initial number in the repository, before the application of data filters and restrictions.

| ID | E.P. | Platform | n. pats |
| --- | --- | --- | --- |
| TCGA-PRAD | PFS | Illumina HiSeq 2000 (mRNA)<br>Illumina HumanMethylation450 BeadChip (methyl)<br>Reverse Phase Protein Array (RPPA) Expression (proteomics) | 495 |
| MSKCC | DFS | Affymetrix Human Exon 1.0 ST arrays | 131 |
| GSE70769 | BCR | Illumina HumanHT-12 V4.0 expression beadchip | 92 |
| GSE54460 | OS | Human 6k Transcriptionally Informative Gene Panel for DASL | 106 |
| GSE46602 | BCR | Affymetrix Human Genome U133 Plus 2.0 Array | 36 |
| GSE53922 | OS | Illumina HumanWG-6 v3.0 expression beadchip | 112 |
| GSE84042 | BCR | Illumina HumanMethylation450 BeadChip<br>Affymetrix Human Transcriptome Array 2.0<br>Affymetrix Human Gene 2.0 ST Array | 160 |
| GSE37199 | HR-LR | Affymetrix Human Genome U133 Plus 2.0 Array | 107 |

**Table 10.**
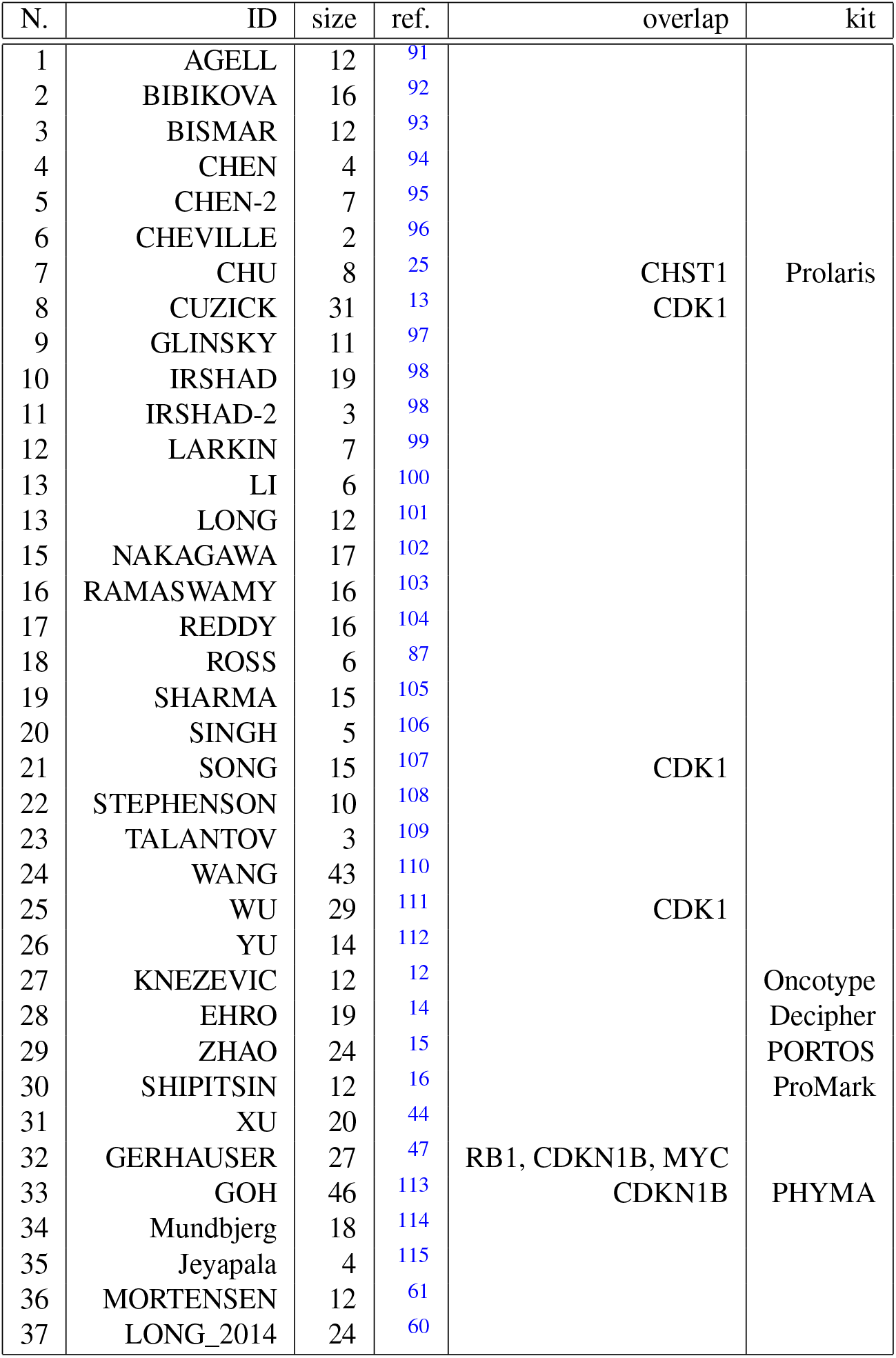
Composition comparison of the CVN fingerprints with published fingerprints in prostate cancer. The table lists progressive number (N.), the fingerprint ID by name of the first author (ID), the published fingerprint size (size), a bibliographical reference (ref.), the genes in common with any of our 7 fingerprints (overlap), and a note of eventual commercial name of an associated prognostic kit (kit). The intersection takes into account gene name aliases as reported by GeneCards - The Human Gene Database https://www.genecards.org/

**Table 11.** Cancer-related annotations for the genes in the pool of seven fingerprints selected by CVN. The table reports the fingerprint identifier (fp), the unique genes in the fingerprint, the cancer types for which the gene has prognostic power for Overall Survival, according to The Human Protein Atlas database - https://www.proteinatlas.org - (Protein Atlas), for non-coding genes the molecular type (note), the most stringent annotation of the gene in the COSMIC (Catalogue Of Somatic Mutations In Cancer) database - https://cancer.sanger.ac.uk/cosmic/ (COSMIC). Note that fp30 shares many genes with fp12, which are reported once.

| fp | gene | Protein Atlas | note | COSMIC |
| --- | --- | --- | --- | --- |
| fp0 | CHST1 | renal, liver |  | no |
|  | GHRL | - |  | mouse gene |
|  | MAK | - |  | no |
|  | RAB11FIP4 | renal,stomach,colorectal |  | no |
|  | RPEL1 | - |  | no |
|  | ZEB1 | renal |  | census tier 2 |
| fp1 | ASH1L-AS1 | renal |  | (ASH1L) mouse gene |
|  | PIFO | renal, pancreatic |  | mouse gene |
|  | DBN1 | renal |  | no |
|  | HRSP12 (RIDA) | liver |  | mouse gene |
|  | MAFG | liver, endometrial |  | no |
|  | SNORA18 |  | snRNA |  |
|  | TRIM65 | liver, renal |  | mouse gene |
| fp14 | CDH1 | renal |  | hallmark |
|  | DIABLO | renal |  |  |
|  | EGFR | urothelial |  | hallmark |
|  | GAB2 | renal |  | mouse gene |
|  | PRKCA | - |  | mouse gene |
|  | RPS6KB1 | renal |  | mouse gene |
| fp12 | CDKN1B | liver, colorectal,renal |  | hallmark |
|  | MAPK9 | colorectal |  | no |
|  | MYC | renal,urothelial,ovarian |  | hallmark |
|  | NDRG1 | liver, renal |  | hallmark |
|  | NF2 | renal |  | hallmark |
|  | RB1 | ovarian |  | hallmark |
|  | SCD | renal, urothelial |  | no |
| fp30 | CDK1 | renal,liver,pancreatic,lung, cervical |  | no |
|  | CLDN7 | renal, thyroid, stomach |  | mouse gene |
| fp20 | BAK1 | renal, endometrial, liver, lung |  | no |
|  | PTCHD4 | renal |  | no |
|  | FANCC | renal |  | census tier 1 |
|  | FBRSL1 | renal, urothelial, prostate |  | no |
|  | OMP | - |  | no |
|  | SULT1C3 | - |  | no |
| fp37 | CCR10 | - |  | no |
|  | NRN1 | renal cancer |  | no |
|  | NPR3 | renal cancer |  | no |
|  | C14orf23 |  | LINC |  |
|  | ATXN7L1 | - |  | no |

We searched the COSMIC (Catalogue Of Somatic Mutations In Cancer) database^5^ for annotations (see Table 11), and we record seven fingerprint genes annotated as cancer “hallmark genes”, nine annotated as “mouse genes”^6^, and two annotated as “census genes”^7^. The hallmarks are concentrated in fp12 and fp30, which share many genes, and fp14. Mouse and census genes are abundant in fp0 and fp1.

#### 2.5.2 Fingerprint fp0

Fingerprint fp0 consists of six genes, namely: CHST1, GHRL, MAK, RAB11FIP4, RPEL1, and ZEB1. Of these, four (GHRL, MAK, ZEB1, and Rab11-FIP4) have been studied in cell and animal models of PRC^21,22,23,24^, one (CHST1) has been included in a published fingerprint^25^, while one (RPEL1) does not appear to have been a focus of study in relation to PRC.

Ye et al.^21^ study PC3 cell lines and *in vivo* mouse models of PRC showing that GHRL mRNA gene expressions and protein levels are increased in invasive PRC. Live imaging in mice models showed that there were different signal intensities of GHRL/GHSR peptide binding in tumor areas with different invasiveness.

Wang et al.^22^ report that MAK dual phosphorylation of the conserved TDY motif is required for MAK kinase activation and that this phosphorylation displays a dynamic pattern during the cell cycle. MAK also acts as a negative regulatory kinase of *APC/C*^*CDH*1^. Interestingly, the CDH1 gene also emerges from our prognostic fingerprint selection process.

Orellana et al.^24^ report that ZEB1 expression correlates with Gleason score in PRC samples and that expression of ZEB1 regulates epithelial–mesenchymal transition and malignant characteristics in PRC cell lines.

He et al.^23^ knocked-out Rab11-FIP4 in PANC-1 pancreatic cancer cells using the CRISPR/Cas9 system and found that this alteration inhibited cell growth, invasion, and metastasis, and arrested cell cycle progression, but did not alter apoptosis.

#### 2.5.3 Fingerprint fp1

Fingerprint fp1 has 7 genes, namely: ASH1L-AS1, C1orf88, DBN1, HRSP12, MAFG, SNORA18, and TRIM65. Two of these have been studied for their role in cancer development. Ye et al.^26^ performed rescue assays on PRC cell lines showing that MAFG may play a key role in facilitating PRC progression. Wang et al.^27^ performed knockdown of TRIM65 in two lung cancer cell lines, SPC-A-1 and NCI-H358, resulting in a significant reduction in cell proliferation, migration, invasion, and adhesion with an increase in G0-G1 phase arrest and apoptosis.

#### 2.5.4 Fingerprint fp12

Fingerprint fp12 has seven genes/proteins, namely: CDKN1B, MAPK9, MYC, NDRG1, NF2, RB1, and SCD. All of them are known to be involved in PRC progression from cell lines and *in vivo* animal models.

Sirma et al.^28^ use large tissue microarray (TMA) from 4699 hormone naive prostate cancers, obtained from patients who had undergone radical prostatectomy, and showed that the loss of CDKN1B/p27 expression was correlated with ERG fusion-negative tumors. The authors however could not identify a direct effect of p27 expression on prostate cancer phenotype or patient prognosis.

Xu et al.^29^ review the role of the JNK family (including JNK1, JNK2 (alias MAPK9), and JNK3) in prostate cancer progression The JNK family has been shown to activate multiple substrates to modulate apoptosis, proliferation, tumorigenesis, and inflammation in response to various stimuli, with emerging evidence indicating the significant roles of the JNK family and androgen receptor in prostate cancer development.

Koh et al.^30^ review a series of recent studies indicating that MYC appears to be activated at the earliest phases of prostate cancer (e.g., in tumor-initiating cells) in prostatic intraepithelial neoplasia, a key precursor lesion to invasive prostatic adenocarcinoma. This phenomenon is evident also in genetically engineered mouse models.

Sharma et al.^31^ report experimental and clinical evidence suggesting that N-myc downregulated gene 1 (NDRG1) functions as a suppressor of prostate cancer metastasis. Their conclusions are based on a three-dimensional invasion assay and an *in vivo* metastasis assay for human prostate xenografts.

Several studies show that inactivation of NF2 contributes to the progression of cancer towards a highly invasive and chemoresistant state^32^.

Han et al.^33^ engineered RB1-depleted C4-2 cell and showed that RB1 silencing resulted in significantly increased cell proliferation and decreased growth response to enzalutamide, a potent AR antagonist.

Fritz et al.^34^ show that pharmacological inhibition of SCD1 activity limits lipid synthesis and results in decreased proliferation of both androgen-sensitive and androgen-resistant PC cells.

#### 2.5.5 Fingerprint fp14

Fingerprint fp14 had 6 genes/proteins, namely: CDH1, DIABLO, EGFR, GAB2, PRKCA, and RPS6KB1. For all of them, there is evidence of their involvement in key cancer development processes.

E-Cadherin (CDH1) is linked with low-penetrance susceptibility that is important in the development of cancer^35^

Kim et al.^36^ demonstrate that the interaction between Smac/DIABLO and Survivin in the nucleus is an important step for suppressing the anti-apoptotic function of Survivin in Docetaxel-induced apoptosis for DU145 prostate cancer cells.

Nastaly et al.^37^ indicate EGFR is a stable, EMT-independent, marker of PRC metastasis to rigid organs, in particular bones.

Tanaka et al.^38^ report that activation of protein kinase C (PKC) by phorbol esters or diacylglycerol mimetics induces apoptosis in androgen-dependent prostate cancer cells.

Hussein et al.^39^ report that suppression of ribosomal protein RPS6KB1 by Nexrutine increases the sensitivity of prostate tumors to radiation both in vitro in multiple PRC cell lines and in the Transgenic adenocarcinoma of mouse prostate model (TRAMP).

Quiao et al^40^ use gene chip technology to screen differentially expressed genes in PC-3 human prostate cancer cells following GRB-associated binding protein 2 (GAB2) gene knockdown, and show that GAB2 regulates several key pathways for PRC insurgence and development.

#### 2.5.6 Fingerprint fp30

Fingerprint fp30 consists of six genes/proteins, namely: CDK1, CDKN1B, CLDN7, MYC, NF2, and SCD. All of them are involved in prostate tumor development. Note that many genes of fp30 are shared with Fp12. Two genes specific of this fingerprint are *CDK*1 and *CLDN*7.

Chen et al.^41^ report that increased CDK1 activity is a mechanism for increasing both Androgen Receptor expression and stability in response to low androgen levels in androgen-independent PCas.

Zheng et al.^42^ show that CLDN7 can regulate the expression of a tissue-specific protein, the prostate-specific antigen (PSA), in the LNCaP prostate cancer cell line

#### 2.5.7 Fingerprint Fp20

Fingerprint fp20 consists of BAK1, PTCHD4, FANCC, FBRSL1, OMP, SULT1C3, and CDKN1B. Two genes in fingerprint fp20 are known to have functional associations with PRC development: BAK1 and CDKN1B.

Shi et al.^43^ showed that transfection of synthetic miR-125b stimulates androgen-independent growth of CaP cells and down-regulates the expression of BAK1.

#### 2.5.8 Fingerprint Fp37

Fingerprint fp37 consists of six methylation loci (listed in Table 3). Using Illumina HumanMethylation450 BeadChip annotations we locate the gene most likely affected by the methylation sites in our fingerprint fp37.

The methylation site cg02928644 is annotated in the database http://www.ewascatalog.org (The MRC-IEU catalog of epigenome-wide association studies) as linked to sex and age, but lacks association with a protein-coding gene.

The other five methylation sites of fp37 are associated with the genes CCR10, NRN1, NPR3, C14orf23, and ATXN7L1. Some of these genes have been studied in relation to other types of tumors, however, their role in prostate cancer is not established. These genes do not appear in the list of hub gene drivers compiled by Xu et al.^44^ for prostate adenocarcinoma. See Lam et al.^45^ for a comprehensive listing of methylation-based biomarkers in prostate cancer.

### 2.6 Overlap with published multi-gene signatures

In Table 10 we list 37 published multi-gene fingerprints developed for prostate cancer (for uses ranging from prognostic to predictive) and we compare them for overlaps with our seven signatures (in Table 3). For fingerprint fp37 we use the genes associated with (closest to) the methylation sites. The published fingerprints have been selected using a comprehensive listing by Manjang et al.^46^ by retaining fingerprints of size comparable with ours (i.e. *<* 100 genes), that are specific for prostate cancer. Moreover, we added fingerprints associated with commercial prognostic/predictive kits. The overlaps for most of the published fingerprints are minimal: gene CDK1 is shared with 3 published fingerprints and gene CHST1 with one. Interestingly, we found three genes (RB1, CDKN1B, MYC) overlapping with the 27-genes fingerprint used by Gerhauser et al.^47^ to identify early onset prostate cancer.

### 2.7 Performance on clinical stratifications

Rodriguez et al.^48^ survey over 20 pre-treatment predictive models using various combinations of the three classical prognostic factors (PSA level, tumor stage, and Gleason Score). We have selected two of these stratification methods: one due to D’Amico et al.^49^ and the NICE (National Institute for Health and Clinical Excellence)^8^ criterion^50^. The two methods differ essentially in the thresholds for discriminating the Intermediate Risk (IR) class from the High Risk class (HR). As almost all patients from the TCGA cohort are at high risk according to both stratification criteria, we report results on the independent cohorts, reporting the predictions with high OR and/or high value of kappa (see Table 7). The statistical significance is almost always attained, except for 3 cases due to the small number of patients involved. Overall CVN-based predictors can stratify well by year the HR patients in both systems. The performance of the CVN-based predictors on the NICE IR class is acceptable, but in general lower than for the HR class.

### 2.8 Random fingerprints

Several authors have noticed that fingerprints obtained by sampling uniformly at random in a pool of genes can have statically significant prognostic performance and sometimes outperform fingerprints obtained with other more elaborated (deterministic, or randomized) methodologies^51,11,46^. The methodology proposed in these studies is oblivious to the model-selection phase used in the determination of the competing multi-gene fingerprints. Here instead we apply a novel comparison methodology against randomly generated fingerprints that is sensitive to the model-selection phase so that random fingerprints and competing fingerprints are treated evenly. We report in Table 8 the performance of the selected random fingerprint out of 100 randomly generated gene fingerprints, using the same model selection methodology we used to attain the fingerprints listed in Table 3, involving both Pareto-based and Ng-based model selection. We find that in two cases (fp14, fp37) the random analog does not attain statistical significance neither in OR nor AUC p-values. In one case (fp0) the random analog attains statistical significance but has quite low performance. In four cases(fp1, fp20, fp30, and fp12) the random analogs are statistically significant and attain good performance, in terms of AUC and kappa measures, although they lag for the corresponding OR measures.

### 2.9 Comparisons with Auto-weka predictors

In Table 12 we report the performance of CVN versus the ML methods in the Auto-weka package (version 2.6)^52^ for the Weka ML environment^53^. Following the same protocol in Pellegrini (2021)^5^ we optimize the hyperparameters for Cohen kappa statistics over 27 base classification methods, 10 meta-methods, and two ensemble methods. Moreover, we apply explicitly seven feature selection methods (including no selection). The reported kappa statistics are computed on the predictor trained on the train data and applied to the test data set. The overall outlook of this experiment with prostate cancer data is quite similar to that on breast cancer data reported in Pellegrini (2021)^5^. Over seven data sets corresponding to the seven fingerprints we have discovered, CVN leads in four cases, ties in one, and loses in two. In each case a different Auto-weka algorithm is attaining the top Auto-weka performance, thus making it hard to pinpoint a single winner algorithm in the Auto-weka suite. Keeping experimental differences in mind, we can confirm the conclusion^5^ that CVN has a level of performance at least comparable with existing ML methods. Moreover, as previously noted, CVN is a single easy-to-explain method that allows for a more uniform approach to the PRC prognosis problem over a wide spectrum of clinical conditions.

**Table 12.** Comparative results of CVN and the Autoweka ML environment. The table reports the input file ID (file n.) corresponding to the seven fingerprints in Table 3, the number of patients in the test set (n. pats), the value of Cohen’s kappa for CNV and for Autoweka, along with the algorithm and the feature selection method attaining it. Autoweka is trained on the corresponding training set via ten-fold cross-validation.

| file n. | n. pats | CVN kappa | autoweka kappa | algorithm | feature selection |
| --- | --- | --- | --- | --- | --- |
| 0 | 53 | 0.29 | **0.32** | Bagging | Corr-Ranker |
| 1 | 37 | **0.54** | 0.45 | SGD | Corr-Ranker |
| 12 | 39 | **0.47** | **0.47** | Random Tree | J48-Ranker |
| 14 | 19 | 0.49 | **0.57** | AdaBoost | Cfs-best |
| 30 | 25 | **0.53** | 0.33 | Simple Logistic | All genes |
| 20 | 39 | **0.43** | 0.21 | Lazy LWL | J48-Ranker |
| 37 | 31 | **0.59** | 0.17 | Lazy Ibk | Corr-ranker |

## 3 Methods

### 3.1 Overiew

Figure 9 shows a schematic depiction of the two main software pipelines used to derive the results reported in this work. In this section, we give a summary of the main principles of the Coherent Voting Network paradigm while more algorithmic details are in Pellegrini 2021^5^. Novel algorithmic features described below include a model selection module based on a theory by Andrew Ng for avoiding model overfitting, and the implementation of a bootstrapping module in a train-test setting according to the work by Efron and Tibshirani 1997^18^.

**Figure 9.**
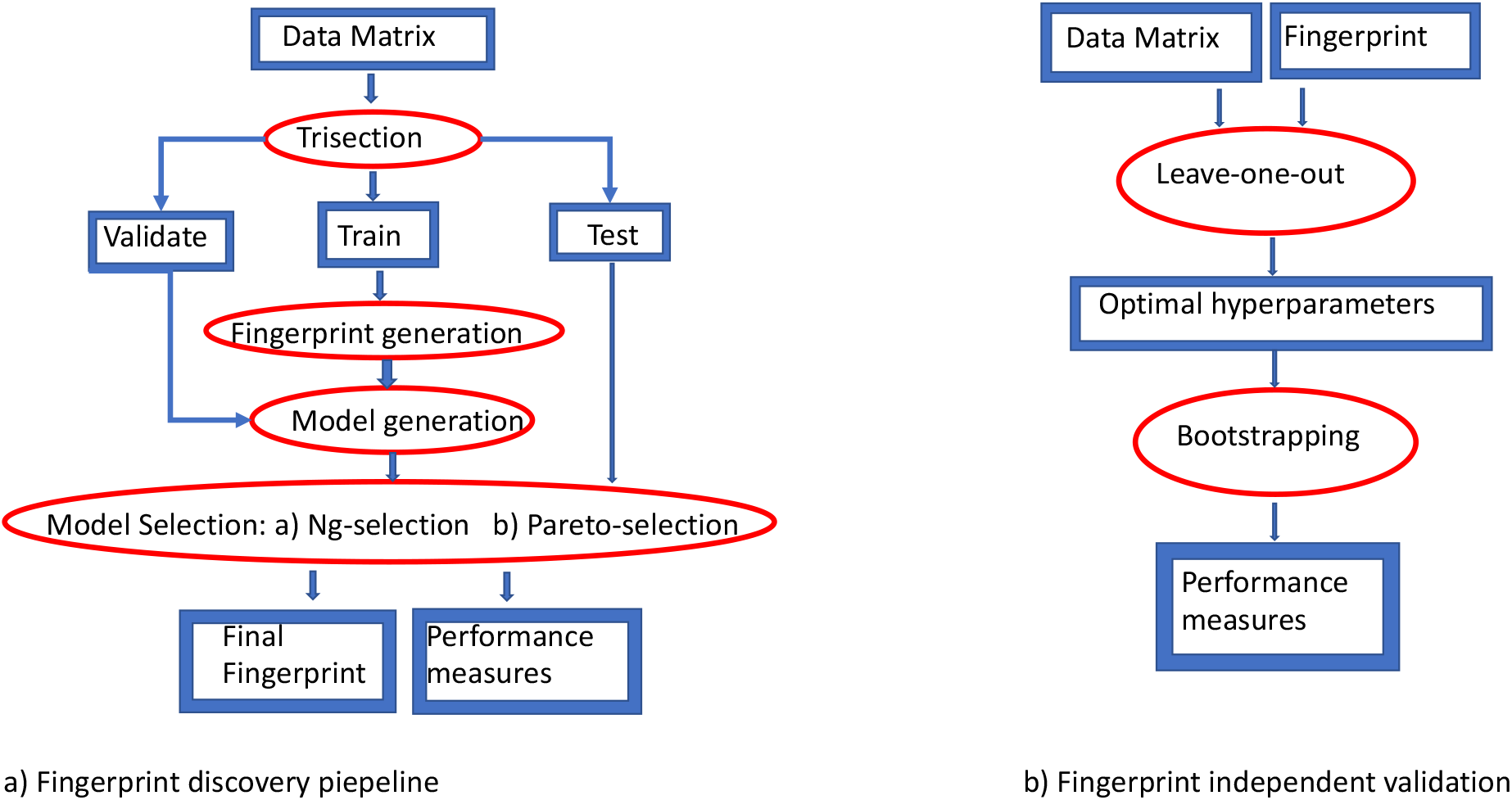
Schematic depiction of the two main software pipelines used in this work. a) Pipeline for fingerprint discovery and evaluation based on a train-validation-test scheme. b) Pipeline for evaluation of a given fingerprint on independent cohorts based on a leave-one-out and bootstrapping scheme.

### 3.2 Coherent Voting Networks

The Coherent Voting Network (CVN) is a supervised learning algorithm introduced by Pellegrini (2021)^5^ and applied to the classification of breast cancer patients into prognostic survival categories (low risk/high risk of overall survival above/below 5 years) after surgical removal of the tumor^5^. The Coherent Voting Network is designed explicitly to uncover non-linear, combinatorial patterns in complex data, within a statistically robust framework. Moreover, the *coherent voting communities* metaphor can be seen as a ‘white box’ approach, providing a certificate justifying the survival prediction for an individual patient, thus facilitating its acceptability in practice, in the vein of explainable Artificial Intelligence.

In a nutshell, CVN can be seen as a generalization of the notion of guilt by association (GbA) in biological networks, where an unlabeled patient node receives a predicted label by collecting the vote of many dense communities of labeled patients and genes to which the unlabeled patient node belongs. The CVN algorithm also seeks a minimal number of genes with the property of allowing a coherent vote of high accuracy on the labeled nodes, and thus such minimal set represents arguably a good candidate fingerprint to be performing well also on predictions for the unlabeled nodes. For further details, we refer the reader to Pellegrini (2021)^5^ and its supplementary materials.

As in many complex ML paradigms, the CVN depends on a number of inner parameters, and thus it is important to do properly both feature selection (i.e. the selection of the fingerprint genes) and hyper-parameter optimization. These two tasks are called together the *model-selection* phase.

The input cohort of patients is split randomly into a training set, a validation set, and a test set (of size roughly 1/2, 1/4 and 1/4). Then we have three phases. In Phase I the CVN is applied to the training set (with full knowledge of the training patient survival labeling) in order to produce a list of candidate gene fingerprints (typically a number between 30 and 60 candidates in our experiments). In phase II, the candidate fingerprints, the training set, and the validation set (with partial knowledge of the patient survival labeling for the validating set) are used together to do model-selection and fix both the fingerprint and the hyper-parameter configuration that minimizes the generalization error (or other performance target measures). Finally, in Phase III we apply the single CVN so built to the test set to measure the effective generalization error. The test set is a set of patients not used in phases I and II, thus unlikely to suffer from overfitting.

We noticed that the standard model selection method suffers from a particular type of overfitting discovered by Andrew Y. Ng^6^ as an effect of having a large number of hypotheses to choose from. we solved this problem by introducing a Pareto stratification^5^ of the models, and by using the notion of a limited and controlled lookup of test data during the model-selection phase (phase II). The lookup of 1 corresponds to the standard model selection, while we considered acceptable also lookup numbers less or equal to 4. Here we effectively attained the prevention of overfitting by using a controlled information leak.

The fingerprints so selected were next further validated in independent cohorts of cancer patients, thus showing that the Pareto-based model selection did perform well empirically.

The main technical contribution of this paper is a new look at the problem of model selection by generalizing and expanding the approach proposed by Andrew Y. Ng^6^, as described in the next section. In practice, we use both the Pareto-based model selection and the Ng-based model selection to attain the results shown in this paper.

### 3.3 Ng-based model selection

In Ng (1997)^6^ it is described the following phenomenon. One has many predictive models (hypothesis) to choose from and uses cross-validation on a pool of validation data in order to select the hypothesis minimizing the cross-validation error, as a representing a hypothesis hopefully minimizing also the generalization error (to be evaluated on a different independent testing set drawn from the same distribution). Ng shows that, when the number of hypotheses to choose from is large, a form of over-fitting occurs so that the hypothesis minimizing the cross-validation error is a poor predictor of the generalization error. Next, an algorithm called LOOCVCV is proposed to cope with this phenomenon^6^. LOOCVCV is based on estimating the number 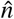 so that choosing the hypothesis with the smallest cross-validation error in a random subset *H*^*′*^ of size 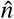 of the initial set *H* of hypotheses has the minimum expected misclassification error. Having the estimate of 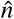, this value is then used in an index-scaling approach to select one of the hypotheses in a ranked list (by cross-validation error) of the initial *H* hypotheses.

We modify and generalize the LOOCVCV method in four aspects:

1. we extend this paradigm to optimize the expected generalization value of functions different from the generalization error, in particular to the Cohen’s kappa measure (and variations of it).
2. we simplify the handling of ties in the ranking of *H* by using lexicographic sorting of the value of a function paired with the index of the hypothesis.
3. we skip the index-scaling approach to the hypothesis selection by recording in the computation process of the estimate of 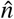, the hypothesis having the largest (smallest) contribution/effect when we aim at maximizing (minimizing) a target function.
4. probabilities of events are computed exactly via binomial coefficients, not in a quick but approximate fashion^6^.

The presence of possible no-predictions introduces some complications, as the Cohen kappa can be changed in several different ways. We compute four versions of the kappa function differing in the way they handle the no predictions. The first solution is to apply the standard Cohen’s kappa functional just ignoring the no predictions. The second solution is to scale the first solution by the fraction of predictions. The third solution is to apply Gwet’s version of kappa^54^. Finally, we consider also a mixed version that uses the second function for a number of no predictions below 15% and the third version when the number of no predictions is above 15%. These four measures are all in the range [*−*1, +1]. In order to select dynamically one of the four measures, we normalize each of them with respect to their own empirical distribution via a z-score. Among these four functions then we choose the function realizing the largest z-score (i.e. scaled displacement from the respective mean).

### 3.4 Bootstrapping

The independent cohorts we use to validate the chosen fingerprints are smaller than the TCGA-PRAD cohort we used to discover them. Therefore splitting these data into three sets risks producing results lacking statistical significance just due to the small numbers involved. For this reason, we use a different common machine learning paradigm. We use a leave-one-out (LOO) approach to hyper-parameter optimization (now the features - genes- are fixed), and we use bootstrapping to evaluate the quality of the chosen configuration^55^.

Bootstrapping is a very general technique with deep theoretical support and extensive practical applications. In the context of cross-validation, we adopt the formalism by Efron and Tibshirani^18^. In particular, we notice that the formula for the leave-one-out bootstrap error estimation (which is the smoothed version of the standard cross-validation estimation of the prediction error) can be applied to obtain smoothed estimates of any function that is a sum (linear combination) of the single error indicator functions for the elements of the set. Therefore we can make bootstrap estimates of the relevant quantities: TP (True Positive), FP (False Positive), TN (True Negative), FN (False Negative), and NP (No Prediction). From these values, we can compute estimates of the bootstrapped odds ratio and kappa. Note that the AUC does not have the required functional form for the application of the theory^18^. For AUC, we proceed as follows. We collect all the prediction maps produced in the bootstrap process and for each patient in the input set and we build a consensus prediction that is the majority of the predictions in the collections of bootstrap maps. Finally, we can compute the AUC of the consensus prediction map using the equivalence to the Wilcoxon-Mann-Whitney U-Statistic.

In standard bootstrapping the sampling in a set of *n* items is done by sampling uniformly at random *with replacement m* = *n* times. Most of the bootstrap theories would carry on using a number of samples *m* ≠ *n* (see e.g. Bickel et al.^56^ for the correction to the theories need in this case). Note that the only practical effect of sampling in our context is to partition the input set into an in-set and an out-set For our experiments on pure genomic fingerprints, we use *m* = 3*n* which ensures sufficient variability in the size of the out-sets (used for testing) while ensuring that the in-sets (used for training) are sufficiently stable. For the mixed clinical and genomic fingerprint on TCGA data, we use *m* = 1.38*n* ensuring that the expected size of the test subset of patients is 1/4 of the total in each bootstrap round, thus with a split close to the initial train-validate-test setting. The results in table 5 are obtained for *B* = 200 and are stable with respect to the number *B* of bootstrap iterations.

### 3.5 Discovery cohort and independent validation cohorts

#### 3.5.1 Discovery cohort: TCGA-PRAD

The discovery cohort is the TCGA-PRAD (2018) data set downloaded from cbioportal^9^ (additional clinical data has been obtained from UCSC Xena repository^10^). The procedures for sample selection and processing are described in detail in the paper by Abeshouse et al.^3^ and its Supplementary files. Briefly, surgical resection biospecimens were collected from patients at the participating institutions diagnosed with prostate adenocarcinoma, who had not received prior treatment for their disease (chemotherapy, radiotherapy, or hormonal ablation therapy). The specimens comprise primary tumor tissue, normal solid tissue, and blood-derived normal. Pathology quality control was performed on each tumor and normal tissue specimen from a frozen section slide. Hematoxylin and eosin (H&E) stained sections from each sample were subjected to independent pathology review to confirm that the tumor specimen was histologically consistent with the allowable prostate adenocarcinoma subtypes and the adjacent normal specimen contained no tumor cells. Computational pipelines include batch effect analysis and correction. Note that in our study we use only the primary tumor-tissue data and clinical data. Some technical details of the data acquisition technology are summarized in Table 9. Although TCGA data was not originally collected for survival analysis, ex-post quality control studies by Liu et al.^19^ show that TCGA PRAD data for PFS is of high quality and can be safely used for our purposes.

#### 3.5.2 Independent validation cohorts: tumor-based samples

##### MSKCC

The data set MSKCC (Cancer Cell 2010) has been downloaded from cbioportal. Study data is also deposited in NCBI GEO under accession number GSE21032. Details of the patient selection and data processing are described by Taylor et al.^57^. In summary, a total of 218 tumor samples and 149 matched normal samples were obtained from patients treated with radical prostatectomy at Memorial Sloan-Kettering Cancer Center. All patients provided informed consent and samples were procured and the study was conducted under Memorial Sloan-Kettering Cancer Center Institutional Review Board approval. Clinical and pathologic data were entered and maintained in MSKCC prospective prostate cancer database. After radical prostatectomy, patients were followed with history, physical exam, and serum PSA testing every 3 months for the first year, every 6 months for the second year, and annually thereafter. Biochemical recurrence (BCR) was defined as PSA *≥* 0.2 ng/ml on two occasions. Note that in our study we use only the primary tumor-tissue data and clinical data. Some technical details of the data acquisition technology are summarized in Table 9.

##### GSE70769

Data from the study of Ross-Adams et al.^58^ was obtained from NCBI GEO under accession number GSE70769. Briefly, the discovery cohort comprises 358 fresh frozen samples from 156 men, including 125 primary prostate cancer from radical prostatectomy with matched benign tissue, 64 matched germline genomic DNA, 19 castrate-resistant prostate cancer (CRPC) from channel transurethral resection of the prostate, 13 with matched germline gDNA, and 12 independent benign samples from holmium laser enucleation of the prostate. Samples were prepared as described in Warren et al.^59^. Relative proportions of benign, epithelial, stromal, and tumor cells were determined by consultant histopathologists; samples with *≥* 20% tumor and matched non-tumor cores (when available) were included. In our study, we use only data from the 125 primary prostate cancer and the 19 castrate-resistant prostate cancer (CRPC) cases and clinical data. Some technical details of the data acquisition technology are summarized in Table 9.

##### GSE54460

Data from the study of Long et al.^60^ was obtained from NCBI GEO under accession number GSE54460. We refer to Long et al.^60^ for more details on the patient selection process and the data processing techniques. In brief, this data set comprises RNA samples passing QC analysis from the Atlanta VA Medical Center (AVAMC), the U. Toronto Sunnybrook Research Centre (UT), and the Moffitt Cancer Center (MCC) in Tampa, FL. MCC Prostate cancer cases were men 21 years and older who had surgery (radical prostatectomy) between 1987 and 2003 for their disease at the MCC and had pathologically confirmed primary prostate cancer. AVAMC cases were patients with prostate cancer who underwent radical prostatectomy between 1990 and 2000. University of Toronto (UT) cases were patients with prostate cancer who underwent radical prostatectomy at the Sunnybrook Health Science Center between 1998 and 2006. These patients did not receive neoadjuvant or concomitant hormonal therapy before radical prostatectomy. Some technical details of the data acquisition technology are summarized in Table 9.

##### GSE46602

Data from the study of Mortensen et al.^61^ was obtained from NCBI GEO under accession number GSE46602. Samples for this study were provided by the Aarhus prostate cancer project consisting of all patients undergoing radical prostatectomy at the Dept. of Urology, Aarhus University Hospital from 1995 to 2015. Clinical data were collected prospectively and recurrence status for all patients was updated before inclusion in the study^61^. The prostatectomy specimens were examined by a trained pathologist, the pathological stage was assessed and the Gleason grade of the tumor was determined. Serum PSA was measured prior to surgery by automated immunoassay using DPC Total PSA Immulite and expressed in ng/ml. Follow-up after surgery has been conducted by PSA measurements at 3, 6, and 12 months postoperatively and thereafter biannually. Subsequent biochemical failure was defined as a PSA *≥* 0.2 ng/ml. Biopsies were taken from the surgical specimen and immediately snap frozen. Normal tissue samples were obtained from a different cohort of patients undergoing cystectomy. Note that in our study we use only the primary tumor-tissue data and clinical data. Some technical details of the data acquisition technology are summarized in Table 9.

##### GSE84042

Data from the study of Frazer et al.^62^ was obtained from NCBI GEO under accession GSE84042 (including both methylation and mRNA gene expression data). More details on patient selection and data processing are in Frazer et al.^62^. Briefly, all patients in this cohort underwent either image-guided radiotherapy (IGRT) or radical prostatectomy (RadP), with curative intent for pathologically confirmed prostate cancer and were hormone naive at the time of definitive local therapy. In the IGRT sub-cohort, a single ultrasound-guided needle biopsy was obtained before the start of therapy, All fresh-frozen RadP specimens were obtained from the University Health Network (UHN) Pathology BioBank or from the Genito-Urinary BioBank of the Centre Hospitalier Universitaire de Québec (CHUQ). All patients were of type N0M0 as an entry criterion for this cohort. For IGRT patients, BCR was defined as a rise in PSA concentration of more than 2.0 ng/ml above the nadir (after radiotherapy, PSA levels drop and stabilize at the nadir). For RadP patients, BCR was defined as two consecutive post-RadP PSA measurements of more than 0.2 ng/ml (backdated to the date of the first increase). If a patient has successful salvage radiation therapy, this is not BCR. If PSA continues to rise after radiation therapy, BCR is backdated to time of the first PSA *>* 0.2. If the patient gets other salvage treatment (such as hormones or chemotherapy), this is considered BCR.

#### 3.5.3 Independent validation cohorts: blood-based samples

##### GSE53922

PBMC or plasma samples were obtained from 117 patients with metastatic CRPC who were positive for human leukocyte antigen (HLA)-A2, A24, A3 supertype (A3, A11, A30, A31, and A33), or A26 and enrolled in clinical trials between February 2001 and April 2008 at the participating hospitals in Japan. Whole-genome gene expression profiles of peripheral blood mononuclear cells (PBMCs) in castration-resistant prostate cancer (CRPC) patients was measured before administration of Personalized peptide vaccination. More details on patient selection and data processing are in the study by Araki et al.^63^. Data were obtained from NCBI GEO under accession GSE53922. Some technical details of the data acquisition technology are summarized in Table 9.

##### GSE37199

Data from the study of Olmos et al.^64^ was retrieved from NCBI GEO under accession number GSE37199.

Briefly, whole blood RNA samples were acquired from patients treated at The Royal Marsden Hospital NHS Foundation Trust (Sutton, UK) and The Beatson West of Scotland Cancer Centre (Glasgow, UK) between August 2007 and April 2008. Patients were enrolled in two groups: patients with advanced castration-resistant prostate (ACRPC) cancer; and (2) patients undergoing active surveillance in a prospective research trial (AS). All patients had a histological diagnosis of prostate cancer and provided informed and written consent for these studies, before sample collection. For each patient, 2·5 mL of peripheral venous blood was collected in 5 mL PAXgene tubes. All samples were taken at least 1 month after cessation of any prostate cancer therapy. Additionally, blood was collected 1 month after the first sampling in patients who had not yet been started on a new prostate-cancer treatment.

Whole-blood RNA was isolated and purified with the PAXgene Blood RNA Kit according to the manufacturer’s instructions. RNA quality and quantity measures were done with a 2100 Bioanalyzer (Agilent Technologies, Palo Alto, CA, USA) and an ND-1000 spectrophotometer (Thermo Scientific, Newark, DE, USA), respectively.

Some technical details of the data acquisition technology are summarized in Table 9.

As we could not access detailed follow-up data, we take the classification into AS and ACRPC patients as a proxy for the ground truth classification in low-risk high-risk sub-classes for PFS at 2 years. This choice is consistent with the value of the median OS from patients in clusters LPD1 and LPD2, which are rich in ACRPC patients with survival below 25 months^64^.

## 4 Discussion

Research in prognostic predictions, in general, and for prostate cancer, in particular, is a vast subject with implications from several areas of biology and medicine, therefore here we comment on the relationship of our work with some issues arising in the relevant literature. Each issue is introduced by a short heading.

### Role of AI and ML in biomarker discovery

Alarcón-Zendejas et al.^65^ and Goldenberg et al.^66^ review recent advances in biomarker discovery for prostate cancer, indicating ML-based and AI-based approaches as opening a new dimension to research and opportunities for transferring new computational techniques in clinical practice in this area. In our work, we push this view by extending the novel ML paradigm of the Coherent Voting Networks (CVN) with improved model selection techniques, and by applying it to the challenging problem of the prognosis of prostate cancer at a fine time granularity (year-to-year).

### Prognosis based on gene expression and proteomic data

We use mainly mRNA gene expression data sets obtained via high throughput assays as the primary source for prognostic biomarker discovery and validation. This technology is now mature and, over time, data on many cohorts of patients have become publicly available. The results on mRNA-based fingerprints appear to be robust w.r.t the specific technology used for measuring mRNA levels of expression. Interestingly, some of the best results we report are obtained from proteomic data obtained with Reverse-Phase Protein microArrays (rppa) assays^67^. Such proteomic data, although less abundant than mRNA expression data may have the advantage of representing a more accurate snapshot of the cells biological processes. In our study, we have derived two fingerprints from mRNA data, three from proteomic data, one from mixed mRNA and proteomic data, one from methylation data, and one from mixed proteomic and clinical data.

### Role of methylation in cancer

Many studies indicate that changes in DNA methylation contributes to cancer development and regulation. Cancers characteristically display extensive hypomethylation of DNA repeats as well as frequent focal DNA hypermethylation^68,69^. Toth et al.^70^ attain good prognostic performance with a Random Forest algorithm, to discriminate patients according to eventual recurrence-free survival as an outcome, measured by PSA levels. However, the model they describe requires input from a large number of methylation sites (402 differentially methylated sites). Our methylation-based fingerprint comprises just six methylation loci with performance validated in the independent GSE84042 methylation data set.

### MicroRNA, microbiome, and Copy Number Alterations

MicroRNA have been investigated as potential biomarkers for PRC prognosis as they can be derived also from liquid biopsies^71^, although the majority of studies still use tissue-derived microRNA^72^. Our experiments with microRNA data from the TCGA-PRAD cohort did produce fingerprints with statistically significant but suboptimal performance (data not shown) vs. those obtained via mRNA, rppa, and methylation data. Similarly, statistically significant but suboptimal results were obtained with TCGA-PRAD CNA and microbiome data (data not shown).

Smith and Sheltzer^73^ study the prognostic power of CNA in several cancer types, including prostate cancer, focusing on alterations of known driver genes. They used Cox proportional hazards analysis, concluding that very few mutations were significantly associated with patient outcomes. Their analyses suggested that, in general, cancer driver gene mutations lacked significant patient stratification power. Our results on the TCGA-PRAD CNA are consistent with these findings^73^.

### Prognostic signatures through tissue classification

In our study, we aim at predicting individual prognostic high-risk/low-risk stratification of patients along yearly time-frames in the first 5 years post-surgery/biopsy. Another form of prognostic study aims at a classification of the tumor tissues into sub-types, and then at using this information to derive broad prognostic indications. For example, Dhanasekaran et al.^74^ study the patterns of differential expressed genes in normal adjacent prostate tissue (NAP), benign prostatic hyperlasia (BPH), localized prostate cancer, and metastatic, hormone-refractory prostate cancer, using unsupervised hierarchical clustering. Among the genes cited^74^ as strongly correlated with the above classification, we find two genes (MYC and CDH1) present also in our fingerprints. Rhodes et al.^75^ produced a list of genes consistently up-regulated or down-regulated in several cohorts of prostate cancer patients with clinically localized prostate cancer versus benign prostate tissue. In this list, we find MYC but no other gene in our fingerprint. We infer that, in all likelihood, our fingerprints do not target the known PRC subtypes *per se*, but, instead, aim directly at the relevant biological process in tumors’ development.

### Prognosis based on clinical and histological data

Historically, histological and clinical parameters have been extensively studied in order to provide effective prognostic stratification of PRC patients. This line of research is now being supplemented with AI-based techniques. For example, Guinney et al.^76^ recently used crowdsourced challenges to improve prostate cancer prognostic models based on open clinical trial data, including 150 curated clinical variables, within the DREAM initiatives (Dialogue for Reverse Engineering Assessments and Methods). A hybrid approach is using genomic profiling to reduce the technical and subjective variability in the estimation of well-known clinical/histological parameters. For example, Wang et al.^77^ initially identify the candidate genes related to the Gleason score, then these genes are used to construct a LASSO Cox regression prognostic analysis model based on a 3 genes fingerprint (CDC45, ESPL1, and RAD54L). In this study we have explored predictors composed of mixed clinical and omic feature, finding good and performance, confirmed in independent cohorts, for a fingerprint composed of three well known clinical parameters (PSA, Gleason primary score, and tumor stage) and expression levels for NF2 and CDKN1B. Interestingly these three clinical parameters were not pre-determined, but emerged from a pool of 24 clinical features. Moreover, the fact that these three clinical features are already routinely collected in practice, implies that just two additional ‘omic’ expression measurements need to be collected (possibly by RT-PCR). Integration of clinical and genomic fingerprints has been shown to be beneficial also for the Decipher fingerprint^78^.

### Role of therapies

In our discovery cohort TCGA-PRAD, no patient received neo-adjuvant therapies prior to surgery/biopsy. About a quarter of the patients has a record of some treatment after surgery (radiation or pharmacological), which may have been administered after monitoring revealed the progress of the disease. Since we aim at predicting the duration of progression-free survival (PFS), we did not stratify the patients into treatment classes. Moreover, note that our study is retrospective, and the effect of personalized therapeutic choices can be detected more reliably within randomized clinical trials specifically designed for this objective^79^.

### Multi-gene prognostic tests in clinical practice and guidelines

Beyer et al.^80,81^ recently compiled a systematic review of diagnostic and prognostic biomarkers in prostate cancer, with emphasis on those likely to progress towards clinical practice. Our multi-gene biomarker fingerprints may be useful within the prostate cancer management work-flow as a PRC risk stratification decision point, following a prostate biopsy/surgery, thus we can hypothesize a potential future use akin to that of the current kits such as Promark, Oncotype Dx, Prolaris, and Decipher^82,83^.

### Cross-cancer diagnostic profiles

Zhou et al.^84^ use the transcriptome profiles of 2180 samples with ovarian (OV), prostate (PRAD), and breast (BRCA) cancer tumors from The Cancer Genome Atlas (TCGA) database to train a deep neural network for a more precise diagnosis of solid tumor vs. adjacent normal tissue. It could be interesting to explore whether CVN may improve the discrimination of normal vs tumor by using a cross-cancer approach. Cross-cancer approaches however do not appear to be proper for prognostic uses, since different tumor types may have markedly distinct progression patterns.

### Multi-omic signatures

Fraser et al.^62^ study in-depth the class of localized, non-indolent prostate cancer and propose a multi-modal pool of biomarkers to predict disease relapse as indicated by BCR (this signature includes clinical, gene expression, methylation sites, SNV, and CNA). Interestingly, their method was effective in predicting eventual relapse with AUC 0.83 (See figure 10 (h)^62^). However, when it was applied to detect early relapse (relapse by month 18) it did not perform well (log-rank test p=0.14) (See figure 10 (g)^62^). In contrast, our signatures are effective within the first 2 to 5 years since surgery/biopsy, with 1-year resolution. Most of our fingerprints are composed of one molecular type, except fp20 which is composed of two, and fp160 composed of clinical and genomic (rppa) markers. Several recent studies have focused their attention on providing refined risk stratifications in the early years after primary treatment. Fu et al. 2021^85^ propose an 18-genes genomic fingerprint for prediction of recurrence with AUC performance values of 0.747, 0.827, and 0.851 respectively after 1-, 3-, and 5-years from surgery in the GSE46602 independent cohort. Zhou et al 2021^86^ report prognostic accuracies for 3- and 5-year BCR-free survival of AUC 0.68 and 0.713, respectively, for a 26-patient independent cohort. Results reported in Tables 5 and 6 show that some of the fingerprints reported in this study may attain higher AUC values with shorter fingerprints.

### Tumor tissue vs liquid biopsies

Blood samples have several advantages with respect to tumor tissue samples as biospecimen of choice for prognostic purposes, and several blood-based prognostic signatures have been proposed for prostate cancer.^64,87,63,88^. In particular issues relative to PRC multiclonality and inter-tumor heterogeneity may limit the use of tissue biopsies as a source of reliable prognostic tests^89^. These issues may be mitigated in blood samples. We tested our fingerprints on two independent cohorts with data from blood samples (GSEGSE53922 and GSE37199) and we found that one fingerprint (fp20) retains prognostic power also in both of these cohorts, although with a higher percentage of no predictions. As the biological and transcriptional interplay of primary prostate adenocarcinoma with eventual bone metastasis affecting several components of blood is complex and not well-understood^64,87^, we expect that better results may be obtained by using blood samples (and/or its components, e.g. extracellular vescicles, serum, PBMC and CTC) directly as the target for the biomarkers discovery phase. We leave this task for future research^90^.

### Metastatic Castration-Resistant Prostate Cancer

One of our independent cohorts (GSE53922) is composed mainly of patients at the stage of Metastatic Castration-Resistant Prostate Cancer (mCRPC). We have found that fingerprints fp14, fp20, and fp30 are prognostic with good performance also for this sub-class of PRC patients, although, in this case, further data need to be analyzed to confirm this finding. For fp20 we also have as partial support the result on cohort GSE37199 where fp20 can discriminate CRPC from the indolent form of local PRC.

### Role of the pool of selected genes in cancer progression

Many of the genes in the seven fingerprints we have selected have been studied individually for their role in cancer (of any type), and they affect functionally important cancer biological processes, as determined via knock-out experiments in cell lines and/or animal models of cancer. In some cases, their gene expression is directly modulated by a microRNA with an important role in cancer progression. Although this is not yet sufficient to establish causal relationships between the expression of these genes and tumor development, it is in our opinion, a good stepping stone towards a more complex type of analysis that integrates bio-networks and causality relationships more explicitly in the model.

### Limitations of the current CVN approach

The main limitation in the current state of the CVN methodology is that the biomarker discovery phase is based on the trisection of the discovery cohort into training, validation, and testing sets (for this study roughly half, one quarter, and one quarter, respectively), while the performance of the selected model can be measured reliably only on the testing set. Thus the size of the discovery cohort needs to be rather large in order for the testing set to be sufficient to attain statistical significance. It is an open line of research to extend the CVN method to reach statistical robustness with fewer initial samples.

## 5 Conclusions

This report has two main contributions. From the methodological point of view, we have extended the CVN (Coherent Voting Network) paradigm by providing a novel robust model selection technique to overcome the danger of overfitting inspired by a method of Andrew Ng. Next, we apply the augmented CVN methodology to tackle the problem of stratifying prostate cancer patients in risk classes (for adverse events within 2 to 5 years from surgery/biopsy). We provide several candidate genomic fingerprints to cover different time-frames at a 1-year resolution and usage of different omic (mRNA, rppa, and methylation) and clinical data. These multi-markers fingerprints can help in deciding the monitoring regime to be applied to prostate cancer patients, within an established clinical decision process. Many of the biomarkers in our pool of genes are known cancer hallmark genes or they are shown functionally involved in cancer using animal models or cell lines. The proposed fingerprints appear to be robust in tests with several independent cohorts.

## Data Availability

Data availability
Data supporting the findings of this study are available from the Github repository
https://github.com/MarcoPellegriniCNR/Coherent-Voting-Network-for-PRC-prognosis.
Code availability
Custom software and code availability is to be agreed via licensing contracts with the National Research Council of Italy.

## 6 Ethical statement

Patients were not directly involved in the study.

## 7 Data availability

Data supporting the findings of this study are available from the Github repository https://github.com/MarcoPellegriniCNR/Coherent-Voting-Network-for-PRC-prognosis.

## 8 Code availability

Custom software and code availability is to be agreed via licensing contracts with the National Research Council of Italy.

## B Contributions

M.P. is the sole author of this publication in all its aspects. Corresponding author: Marco Pellegrini.

## Footnotes

1 https://www.wcrf.org/cancer-trends/prostate-cancer-statistics/

2 https://ecis.jrc.ec.europa.eu

3 https://www.cancer.gov/about-nci/organization/ccg/research/structural-genomics/tcga

4 https://www.proteinatlas.org

5 https://cancer.sanger.ac.uk/cosmic/

6 Mouse genes are listed in the Candidate Cancer Gene Database^20^ recording functional effects in cancer mutagenesis experiments supporting the designation of the gene as causative in cancer.

7 Census genes possess a documented activity relevant to cancer, along with evidence of mutations in cancer that change the activity of the gene product in a way that promotes oncogenic transformation.

8 NICE is an executive non-departmental public body of the Department of Health and Social Care in England that publishes guidelines in several areas.

9 https://www.cbioportal.org

10 https://xena.ucsc.edu

